# Implications of progressive lung damage and post-TB sequelae for the health benefits of prompt TB diagnosis in high HIV prevalence settings: a mathematical modeling analysis

**DOI:** 10.1101/2024.08.12.24311198

**Authors:** Melike Hazal Can, Sedona Sweeney, Brian W. Allwood, Susan E. Dorman, Ted Cohen, Nicolas A. Menzies

## Abstract

**Background:** Untreated pulmonary tuberculosis (TB) causes ongoing lung damage, which may persist after treatment. Conventional approaches for assessing TB health effects may not fully capture these mechanisms. We evaluated how TB-associated lung damage and post-TB sequalae affect the lifetime health consequences of TB in high HIV prevalence settings.

**Methods:** We developed a microsimulation model representing dynamic changes in lung function for individuals evaluated for TB in routine clinical settings. We parameterized the model with data for Uganda, Kenya, and South Africa, and estimated lifetime health outcomes under prompt, delayed, and no TB treatment scenarios. We compared results to earlier modelling approaches that omit progressive lung damage and post-TB sequelae.

**Findings:** We estimated 4.6 (95% uncertainty interval 3.4–5.8), 7.2 (5.1–9.6), and 18.0 (15.1–20.0) year reductions in life expectancy due to TB under prompt, delayed, and no treatment scenarios, respectively. Disability-adjusted life years (DALYs) from TB were estimated as 8.3 (6.2–10.6), 12.6 (9.0–17.0), and 27.8 (24.1–30.6) under prompt, delayed, and no treatment scenarios, respectively. Post-TB DALYs represented 9–53% of total DALYs. Modelling approaches that omit progressive lung damage and post-TB sequelae underestimated lifetime health losses of TB by 48–57%, and underestimated the benefits of prompt treatment by 45–64%.

**Interpretation:** Delayed initiation of TB treatment causes greater lung damage and higher mortality risks during and after the disease episode. In settings with co-prevalent TB and HIV, accounting for these factors substantially increased estimates of the lifetime disease burden and life expectancy loss caused by TB.

**Funding:** NIH.

**Research in context:** *Evidence before this study:* Research on long-term sequalae among tuberculosis (TB) survivors has focused on describing the prevalence and nature of these post-TB sequalae, and quantifying their contribution to the overall burden of TB disease. There is limited evidence describing how improvements in TB diagnosis and prompt treatment initiation could affect the overall health losses associated with TB, inclusive of post-TB sequelae. We searched PubMed from database inception until July 19, 2024, with no language restrictions for studies reporting how TB diagnosis and treatment affect post-TB sequelae and lifetime health losses, using the search terms “(tuberculosis OR TB) AND (post-TB OR post-tuberculosis) AND (diagnos*) AND (treat*) AND (model*)”. We retrieved 21 publications based on this search. Of these, one study reported a mathematical modeling approach for estimating lifetime health outcomes and costs by considering the delay in diagnosis, post-TB sequelae, and treatment discontinuation among TB patients in Brazil, but did not simulate changes in lung function during the TB episode.

*Added value of this study:* To our knowledge, this is the first study to investigate the effects of timeliness of TB diagnosis on progressive lung damage and lifetime health outcomes for individuals with TB. To do so, we constructed a mathematical model simulating changes in lung function before, during, and after TB treatment, and simulated multiple counterfactual scenarios for a cohort of individuals presenting to primary health services with undiagnosed TB disease in Uganda, Kenya, and South Africa. We compared the results of this analysis to the estimates produced by earlier modelling approaches that do not represent TB-associated lung damage or post-TB sequelae.

*Implications of all the available evidence:* The results of this analysis showed that post-TB sequelae represent a substantial share of the overall health losses associated with TB, and that better post-TB lung function (resulting from a shorter duration of untreated TB disease) is a major contributor to the overall health benefits of prompt TB diagnosis and treatment. These results are not accurately captured by earlier modelling approaches that did not consider TB-associated lung damage or post-TB sequelae. The findings of this analysis contribute to the evidence base describing how TB interventions can influence lung function dynamics during and after TB disease, and the resulting changes in disability and mortality due to TB.

## Introduction

In 2022, over 10.6 (9.9–11.4) million persons developed tuberculosis (TB), with 1.3 (1.1–1.5) million persons dying with the disease.^1^ While TB can affect multiple organ systems it most commonly affects the lungs, with pulmonary TB (with or without extrapulmonary involvement) representing most global TB cases.

Pulmonary TB progresses through several disease stages, though this process can differ widely between individuals.^2^ Following initial infection *Mycobacterium tuberculosis* (*Mtb*) replicates inside a granuloma within the lungs. An individual progresses to infectious disease when bacteria escape this granuloma, which can happen soon after infection or decades later.^3^ This is marked by damage to local lung tissue due to both bacterial infection as well as the immune response. Initially, individuals may not feel sick, and some will develop chronic subclinical TB or self-cure.^2^ However, many will eventually develop symptoms traditionally associated with TB, including cough, fever, and weight loss. Individuals with symptomatic disease are also likely to experience progressive lung damage, including cavitation and consolidated areas, and involvement of other organs. While some individuals will clear the TB disease without intervention, case fatality is approximately 50% without treatment.^4^ For individuals with impaired immunity disease progression is faster and survival poorer.^5^

Effective TB treatment can arrest the disease process, with symptoms ameliorating over the course of treatment.^6^ However, for many individuals the lungs do not fully heal, particularly if lung damage was extensive before treatment was initiated. This damage can lead to persistent respiratory impairment and elevated risks of conditions such as bronchiectasis, aspergillosis, and lung cancer. Multiple meta-analyses have documented a high prevalence of respiratory impairment among TB survivors, with poorer spirometry results and functional lung health compared to healthy controls.^7,8^ Studies that have adopted quasi-experimental designs to assess the causal effects of TB on subsequent health outcomes have also estimated elevated healthcare utilization and mortality rates among TB survivors compared to matched controls.^9–11^ This collection of persistent sequelae—collectively known as post-TB lung disease (PTLD)—has drawn increasing scientific and policy attention.^12,13^

Mathematical modelling is commonly used to extrapolate from clinical studies to estimate long-term population-level outcomes,^14^ with these analyses reproducing key features of TB epidemiology and natural history.^15,16^ In many published TB models, active disease is represented by a single disease state, sometimes stratified to capture differences in diagnostic test results (e.g., sputum-smear positivity). Moreover, most published models assume complete recovery following TB cure. While simplifications are needed to render analyses tractable, if they produce a biased representation of underlying disease mechanisms, this could produce incorrect policy conclusions.^17^ The potential biases produced by overly-simplified modeling approaches have been demonstrated previously for *Mtb* infection.^16,18^ It is unclear whether simplified models of TB disease progression and post-TB sequelae affect the findings of these analyses.

In this study, we developed a novel model of TB progression, treatment, and long-term health outcomes among individuals with symptomatic TB disease, explicitly modeling the progressive lung damage caused by TB, the recovery of lung function during and after treatment, and how these processes are modified by the presence of comorbid HIV. We used this model to assess the long-term outcomes of TB diagnosis and treatment using detailed data on primary care attendees evaluated for TB in Uganda, Kenya, and South Africa, three settings with high rates of TB and HIV infection. We compared these results to those produced by traditional modelling representations of TB disease, to understand whether the different modeling approaches produce substantively different results.

## Methods

### Study setting and population

We constructed a study cohort representing individuals with symptomatic pulmonary TB presenting at routine healthcare settings. This scenario represents the way most individuals with TB access treatment in low- and middle-income settings. To do so, we obtained data on individuals evaluated for TB in a multicenter diagnostic accuracy study conducted in Uganda, Kenya, and South Africa.^19^ This sample included ≥ 18-year-old primary healthcare attendees who were evaluated for TB based on clinical suspicion. For participants determined to have TB via sputum culture, we extracted data on age, sex, HIV status, and CD4 cell count if HIV positive. We used FEV1 (forced expiratory volume over 1 second, expressed as a percentage of average values in the population) to quantify the extent of TB-related lung damage among individuals presenting for care.^8,20,21^ As study data did not include FEV1 values, we assigned FEV1 values to participants based on published studies describing the distribution of FEV1 among individuals with diagnosed TB. We reweighted trial data to match national TB notifications data by age, sex, and HIV status,^22^ and created an analytic cohort of 10,000 individuals per country. This study was approved by the Institutional Review Board of the Harvard T.H. Chan School of Public Health.

### Modeling approach

We developed a Markov microsimulation model to track individuals in the analytic cohort over time and evaluate their long-term health outcomes under multiple TB diagnosis and treatment scenarios (Figure S1). The model was parameterized to represent each individual in the analytic cohort and updated with a weekly timestep over the lifetime of each modelled individual. Parameter values and sources are given in Table S1, and full model details are given in the Supplement.

#### Lung function dynamics

We used changes in FEV1 to track the evolution of TB-attributable lung damage for individuals with untreated TB disease, during TB treatment, and among individuals surviving the disease episode. In the model, FEV1 was represented as an individual-level characteristic updated on a weekly basis over the remainder of the individual’s lifetime. Changes in FEV1 were determined based on the individual’s TB disease status (untreated TB disease, receiving TB treatment, post-TB period). As a marker of TB disease severity, FEV1 was assumed to affect multiple other model mechanisms and health outcomes, including mortality rates for TB and post-TB, the self-cure rate, and disability weights describing reductions in quality of life due to TB and post-TB.

#### TB self-cure

We assumed some individuals with TB disease would experience self-cure in the absence of treatment. The self-cure rate was calculated as a function of HIV status, CD4 cell count, and FEV1.

#### TB treatment uptake and discontinuation

Individuals with untreated TB disease were assumed to seek diagnosis and treatment at a fixed rate, differentiated by ART status. Individuals receiving TB treatment could complete the regimen (assumed to be the standardized 6-month first-line regimen),^23^ die during treatment, or be lost to follow-up before completion. For individuals lost to follow-up, we assumed that the probability of cure would depend on the timing of discontinuation, with a cure probability of 0% for individuals completing <8 weeks of the regimen.

#### HIV natural history and treatment

The analytic cohort was subdivided by HIV status and receipt of ART. We assumed individuals with undiagnosed HIV would be diagnosed as part of the assessment for TB and initiated on ART. Individuals receiving ART were assumed to discontinue ART at a fixed rate, based on published rates of loss-to-follow-up for routine ART programs. We used CD4 cell count as a marker of HIV-related immune function, and modeled changes in CD4 cell count over time (differentiated by ART status) using approaches used in prior modeling.^24^ CD4 cell count was assumed to influence HIV-related mortality rates, TB-related mortality rates, and TB self-cure rates. For simplicity we did not model future acquisition of HIV infection within the analytic cohort. For individuals receiving both ART and TB treatment we assumed that rates of discontinuation would be dependent: individuals lost to follow-up from one service would also discontinue the other service.

#### Quality of life

We used disability weights to quantify reductions in quality of life attributable to HIV, TB treatment, and post-TB lung damage. HIV disability weights were based on CD4 cell count and ART status. TB and post-TB disability weights were based on FEV1.

#### Mortality rates

All-cause mortality rates were calculated as the sum of background and disease-specific mortality rates. Background mortality rates by sex, age, and country were based on TB- and HIV-deleted life tables. Disease-specific mortality risks from HIV, TB treatment, and post-TB lung damage were calculated as a function of HIV/ART status, CD4 cell count, TB status, TB treatment status, and FEV1 (equation given in Supplement).

### Analytic scenarios

We simulated outcomes for the analytic cohort under three scenarios: ‘*prompt TB treatment’*, as would result from a correct TB diagnosis (assuming a 2-week provider delay before the regimen is initiated), ‘*delayed TB treatment’*, as would result from an incorrect false-negative TB diagnosis, with the possibility that the individual would eventually return to care with ongoing symptoms (4.0 months (95% uncertainty interval: 2.1–8.1) delay for individuals receiving ART, 7.5 months (3.6–15.5) delay for individuals not receiving ART); and ‘*no TB treatment’*, in which the individual never receives TB treatment despite ongoing symptoms. We also examined a ‘*no TB’* counterfactual scenario that simulated future health outcomes for the analytic cohort without TB disease, in order to calculate the incremental difference in life expectancy and other outcomes that is attributable to TB.

### Alternative model specifications

We re-estimated study outcomes under three alternative model specifications, to test whether approaches to modelling lung function and post-TB sequelae affected study conclusions, as compared to earlier modelling approaches.

*Alternative specification 1 (no post-TB sequelae)*: all modelling assumptions and parameter values matched the main analysis, except that individuals with TB were assumed to experience immediate, complete, and permanent recovery of lung function following TB cure.

*Alternative specification 2 (no progressive lung damage)*: all modelling assumptions and parameter values matched the main analysis, except that all parameters affected by progressive lung damage among TB patients (TB-specific mortality rates, self-cure rates, disability weights) were assumed fixed over time. We operationalized this by fixing FEV1 at the average starting value across all simulated individuals, and holding it fixed at this value until TB cure or death.

*Alternative specification 3 (no post-TB sequelae and no progressive lung damage):* all modelling assumptions and parameter values matched the main analysis, except that individuals with TB were assumed to experience no progressive lung damage (as in Alternative specification 2), and individuals cured of TB were assumed to experience immediate, complete, and permanent recovery of lung function (as in Alternative specification 1).

### Outcomes

Outcomes included the fraction dying with TB disease (deaths among individuals with untreated TB disease or while receiving TB treatment), 5-year survival, life expectancy, and disability-adjusted life years (DALYs) lost. To calculate DALYs we summed the non-fatal health losses due to reduced quality-of-life (Years Lived with Disability, YLDs) and health losses from premature mortality (Years of Life Lost, YLLs) due to TB, and also decomposed total DALYs to report the DALYs accruing during the disease episode and the DALYs after TB cure (i.e., from post-TB sequalae). We estimated outcomes for the overall cohort as well as stratified by HIV status.

### Statistical analysis

We created probability distributions quantifying uncertainty in each model parameter, and used 2^nd^-order Monte Carlo simulation to propagate this uncertainty through the analysis.^25^ To do so, we sampled 1,000 values from each probability distribution using a Latin Hypercube Sampling design, and re-estimated outcomes for each of these 1,000 parameter sets. We used the distribution of results to calculate equal-tailed 95% uncertainty intervals, and also calculated partial rank correlation coefficients (PRCCs) describing the sensitivity of study outcomes to uncertainty in each parameter.^26^ The model was programmed in R (v4.3.2) and C++ using the *Rcpp* package (v1.0.12).^27,28^

## Results

### Characteristics of the study cohort

In the overall cohort, 55.1% (661/1199) were male, mean age was 38 years (interquartile range (IQR): 28–45), 49.0% (588/1199) were living with HIV, and of these 84.7% (498/588) were enrolled on ART. Average FEV1 was 63% (IQR: 52%–75%), and for individuals with HIV, the average CD4 cell count was 400 cells per µL (IQR: 150–598). Table 1 describes cohort characteristics by country. Table S2 shows cohort characteristics after reweighting to match the age, sex, and HIV distribution of TB notifications for each country.

**Table 1.**
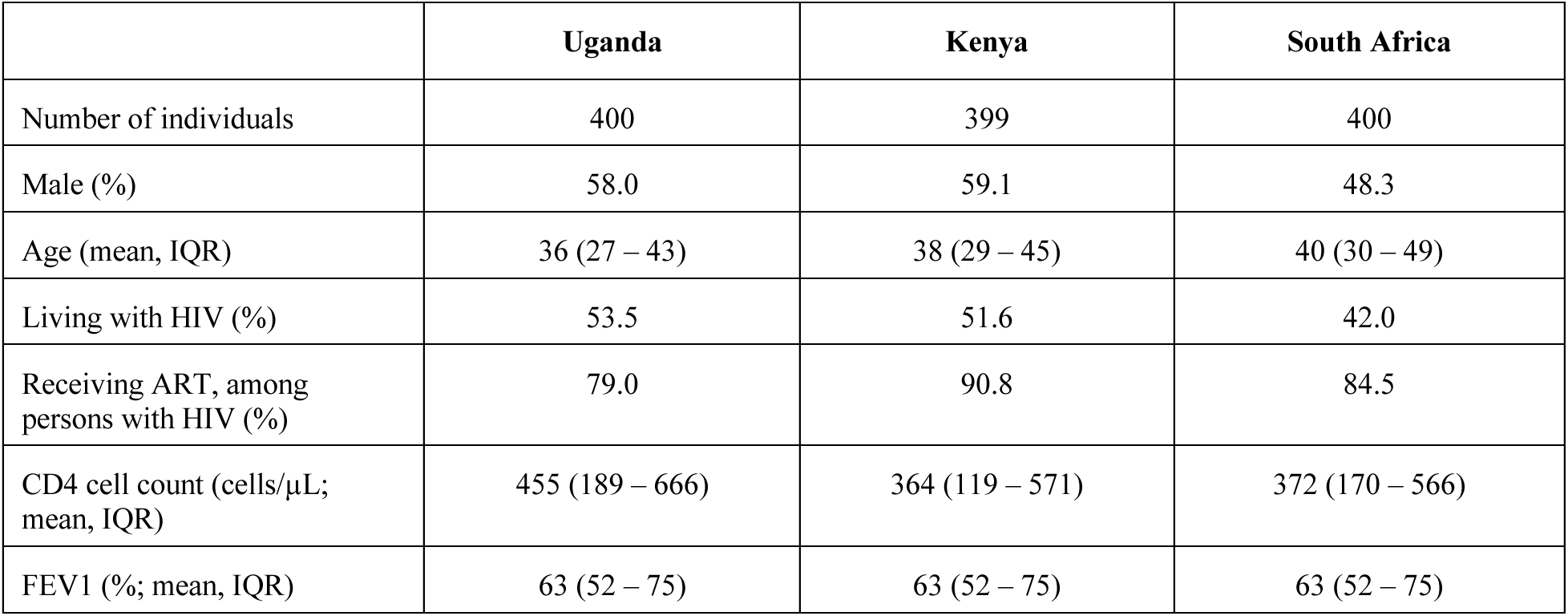
Summary statistics of the study cohort. FEV1: Forced expiratory volume over one second; IQR: Interquartile range; ART: Antiretroviral therapy.

### Mortality trends under each scenario

Figure 1 shows survival curves for persons with and without HIV under each analytic scenario. For individuals without HIV, average survival (from the beginning of the analysis) was 27.9 (95% uncertainty interval: 26.3–29.4) years, 24.9 (21.5–27.5) years, and 12.0 (9.8–14.4) years under prompt treatment, delayed treatment, and no treatment scenarios, respectively. Life expectancy estimates were lower for individuals living with HIV, with average survival of 12.5 (6.6–15.5) years, 10.7 (5.8–13.6) years, and 3.3 (2.2–4.2) years under prompt treatment, delayed treatment, and no treatment scenarios, respectively. Figure S2 shows country-specific survival curves for each scenario.

**Figure 1.**
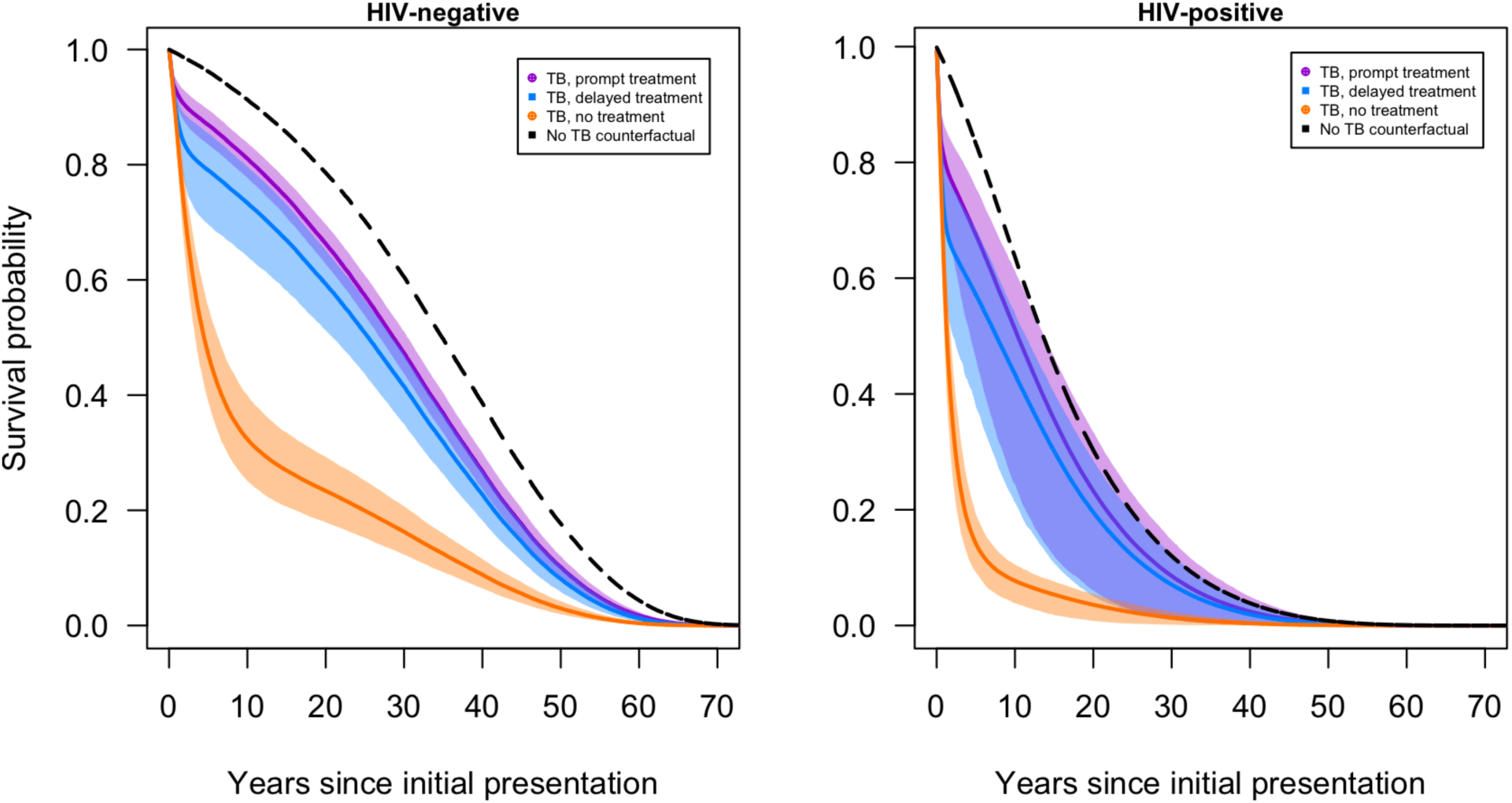
Survival curves for individuals with symptomatic TB under prompt, delayed, and no treatment scenarios.

### Summary health outcomes by scenario

Table 2 shows lifetime health outcomes for each scenario stratified by HIV status, as well as incremental differences in outcomes for each scenario as compared to individuals without TB disease (representing the health losses attributable to TB). For the overall cohort (combining HIV positive and HIV negative individuals), TB was estimated to reduce life expectancy by 5.1 (3.8–6.4), 7.7 (5.5–10.1), and 18.5 (15.5– 20.6) years under prompt treatment, delayed treatment, and no treatment scenarios respectively. Total DALYs attributable to TB were estimated to be 11.4 (8.9–14.2), 17.1 (13.1–22.1), and 37.7 (34.3–40.3) years under prompt treatment, delayed treatment, and no treatment scenarios, respectively. Table 2 also reports incremental differences in each health outcome for delayed treatment and no treatment as compared to prompt treatment (representing the health losses resulting from a missed TB diagnosis).

**Table 2.** Summary health outcomes under prompt, delayed, and no treatment scenarios, stratified by HIV status. ‘DALYs’ = Disability-adjusted life years. Values in parentheses represent 95% uncertainty intervals. * Life expectancy under the no TB counterfactual was 34.0 (33.8, 34.3) years among HIV-negative patients, and 15.8 (7.9, 19.8) years among HIV-positive patients. ** Five-year survival for the no TB counterfactual was 96.2% (95.8%, 96.5%) among HIV-negative patients, and 83.2% (54.4%, 90.5%) among HIV-positive patients. *** As TB-attributable deaths and DALYs are zero under the no TB counterfactual, the absolute value of these outcomes are the same as the difference vs the no TB counterfactual.

|  | HIV-negative |  |  | HIV-positive |  |  |
| --- | --- | --- | --- | --- | --- | --- |
|  | Prompt treatment | Delayed treatment | No treatment | Prompt treatment | Delayed treatment | No treatment |
| **Life expectancy (years) \*** |  |  |  |  |  |  |
| Absolute value | 27.9<br>(26.3, 29.4) | 24.9<br>(21.5, 27.5) | 12.0<br>(9.8, 14.4) | 12.5<br>(6.6, 15.5) | 10.7<br>(5.8, 13.6) | 3.3<br>(2.2, 4.2) |
| Difference vs. no TB counterfactual | 6.2<br>(4.6, 7.7) | 9.2<br>(6.6, 12.5) | 22.0<br>(19.6, 24.2) | 3.3<br>(1.3, 4.9) | 5.2<br>(2.2, 7.9) | 12.5<br>(5.3, 16.2) |
| Difference vs. prompt treatment | Reference | 3.0<br>(1.5, 5.2) | 15.8<br>(14.1, 17.9) | Reference | 1.9<br>(0.7, 3.5) | 9.2<br>(4.1, 11.8) |
| **5-year survival (%) \*\*** |  |  |  |  |  |  |
| Absolute value | 86.9<br>(83.5, 89.6) | 79.0<br>(69.6, 85.4) | 47.1<br>(38.8, 55.3) | 67.5<br>(6.6, 15.5) | 57.2<br>(37.8, 68.4) | 14.2<br>(9.2, 19.4) |
| Difference vs. no TB counterfactual | 29.4<br>(22.3, 36.2) | 40.2<br>(30.5, 51.3) | 74.4<br>(68.3, 79.6) | 19.3<br>(9.3, 26.5) | 29.8<br>(15.5, 42.4) | 69.8<br>(38.6, 79.2) |
| Difference vs. prompt treatment | Reference | 13.9<br>(7.7, 22.5) | 58.6<br>(53.3, 63.7) | Reference | 11.8<br>(5.5, 20.5) | 53.7<br>(30.2, 61.3) |
| **TB-attributable deaths (%)** |  |  |  |  |  |  |
| Absolute value*** | 8.8<br>(6.0, 12.3) | 17.0<br>(10.3, 27.3) | 67.6<br>(60.0, 75.1) | 20.2<br>(14.1, 27.5) | 32.8<br>(22.3, 47.2) | 90.6<br>(87.8, 93.3) |
| Difference vs. prompt treatment | Reference | 8.2<br>(3.8, 15.6) | 58.9<br>(52.6, 65.5) | Reference | 12.6<br>(6.4, 22.2) | 70.4<br>(64.3, 75.2) |
| **DALYs due to TB** |  |  |  |  |  |  |
| Absolute value*** | 11.7<br>(8.6, 14.7) | 17.0<br>(12.2, 23.4) | 35.0<br>(31.5, 38.3) | 11.1<br>(7.9, 14.2) | 17.3<br>(12.1, 23.4) | 39.7<br>(33.9, 41.9) |
| Difference vs. prompt treatment | Reference | 5.4<br>(2.7, 9.2) | 23.4<br>(20.9, 26.1) | Reference | 6.2<br>(3.2, 10.6) | 28.6<br>(24.5, 30.9) |

Compared to prompt TB treatment, life expectancy was reduced by 2.6 (1.5–4.0), and 13.4 (11.2–15.0) years under delayed treatment and no treatment scenarios, and total DALYs attributable to TB increased by 5.7 (3.5–8.6) and 26.3 (23.6–28.5) years.

### Contribution of post-TB sequelae

Figure 2 reports TB-attributable DALYs decomposed according to the form of health loss (reduced quality-of-life vs. premature mortality) and when the health losses occur (TB episode vs. post-TB). These results show the distribution of DALYs to vary by scenario and by HIV status. The proportion of total DALYs accruing during the TB episode increased with greater delays to diagnosis, and was higher for individuals with HIV infection, a consequence of higher TB case fatality in these situations. For the overall cohort, prompt treatment resulted in 52.5% (41.9%–62.5%) of DALYs accruing during the post-TB period (5.4 (4.0–7.2) TB episode DALYs vs. 6.0 (4.1–8.0) post-TB DALYs). For the delayed treatment scenario, the post-TB proportion was 42.7% (31.6%–54.0%) (9.8 (6.9–13.9) TB episode DALYs vs. 7.3 (5.1–10.1) post-TB DALYs), and for the no treatment scenario the post-TB proportion was 9.1% (6.0%–13.3%) (34.3 (30.7–37.2) TB episode DALYs vs. 3.4 (2.3–5.0) post-TB DALYs). Table S3 reports these outcomes stratified by HIV status.

**Figure 2.**
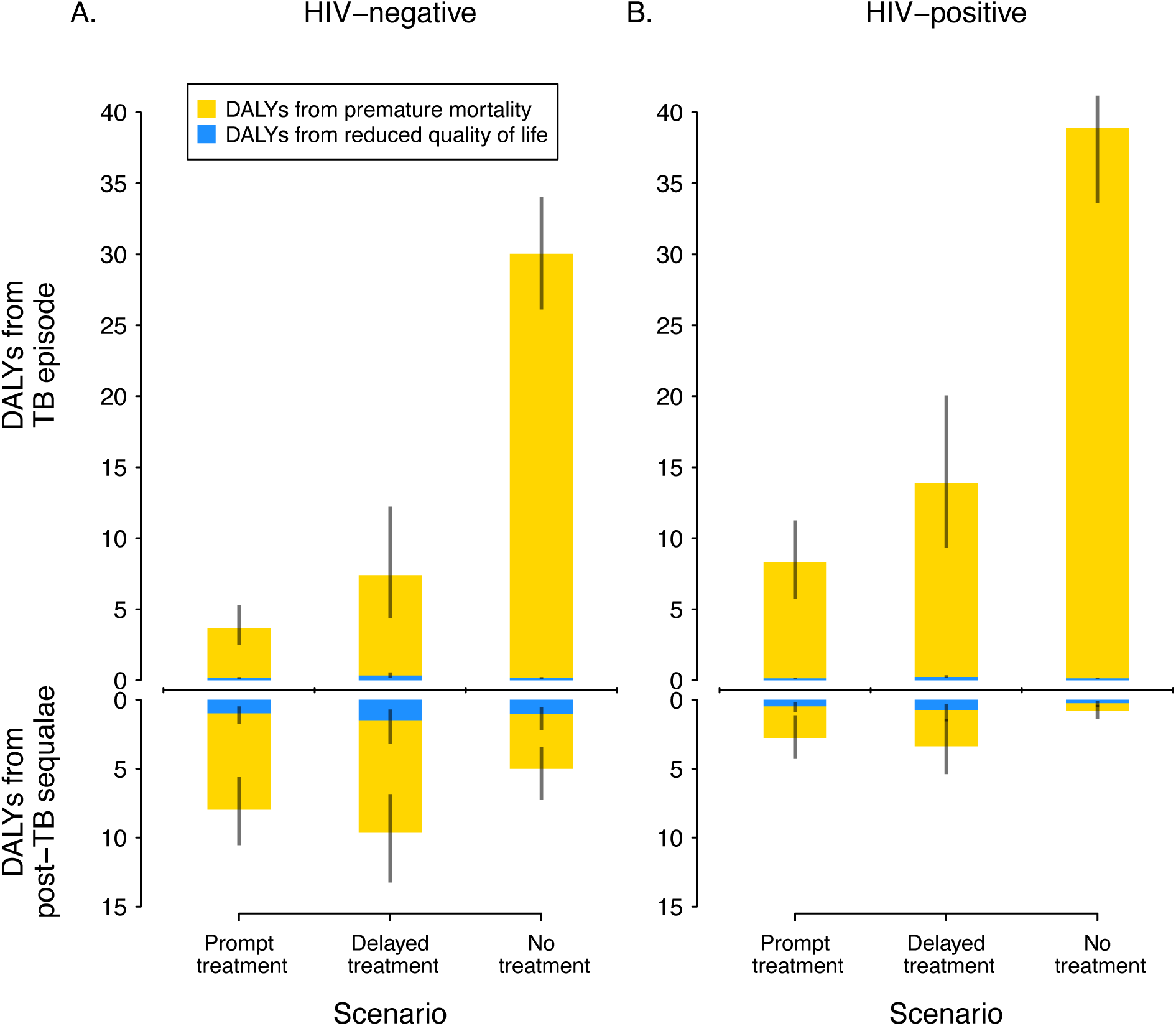
DALYs attributable to TB disease for each scenario by HIV status, stratified by whether DALYs resulted from reduced quality of life or premature mortality, and by whether DALYs occurred during the TB episode or after TB cure.

### Alternative model specifications and sensitivity analyses

The main analysis reports results from a model that accounted for the dynamics of TB-associated lung damage and post-TB sequalae. Table 3 shows how major health outcomes changed under alternative model specifications adopted by earlier modeling studies (i.e., not accounting for progressive TB lung damage or post-TB sequelae). In the specification that omitted post-TB sequelae (alternative scenario 1), estimated life expectancy was longer under each scenario (prompt, delayed, or no treatment) as compared to the main analysis. For these model specifications the estimated reduction in life expectancy caused by TB was substantially lower (8-53% lower) than estimated in the main analysis. For the specification that omitted TB disease progression (alternative scenario 2), estimates of the additional life expectancy loss with delayed or no TB treatment (vs. prompt treatment) were substantially lower (56-67% lower) than estimated in the main analysis. Alternative specification 3 (omitting both post-TB sequelae and TB disease progression) showed the greatest difference from the main analysis, with estimates of TB-attributable reductions in life expectancy 51-55% lower across scenarios, and estimates of the additional life expectancy loss from delayed or no TB treatment (vs. prompt treatment) reduced by 50-62%.

**Table 3.** Summary health outcomes under prompt, delayed, and no treatment scenarios. ‘DALYs’ = Disability-adjusted life years. Values in parentheses represent 95% uncertainty intervals. * As TB-attributable DALYs are zero under the no TB counterfactual, the absolute value of these outcomes are the same as the difference vs the no TB counterfactual.

|  | Prompt treatment |  | Delayed treatment |  | No treatment |  |
| --- | --- | --- | --- | --- | --- | --- |
|  | Estimate | Pct change vs. main analysis | Estimate | Pct change vs. main analysis | Estimate | Pct change vs. main analysis |
| **Life expectancy (years)** |  |  |  |  |  |  |
| *Main analysis* |  |  |  |  |  |  |
| Absolute value | 22.1<br>(19.7, 23.8) | 0% | 19.5<br>(17.0, 21.6) | 0% | 8.8<br>(7.2, 10.5) | 0% |
| Reduction due to TB | 5.1<br>(3.8, 6.4) | 0% | 7.7<br>(5.5, 10.1) | 0% | 18.5<br>(15.5, 20.6) | 0% |
| Reduction compared to prompt treatment | Reference | 0% | 2.6<br>(1.5, 4.0) | 0% | 13.4<br>(11.2, 15.0) | 0% |
| *Alternative specification 1 (no post-TB sequelae)* |  |  |  |  |  |  |
| Absolute value | 24.8<br>(22.4, 26.3) | 12.2%<br>(8.3%, 16.3%) | 22.7<br>(20.0, 24.6) | 16.3%<br>(10.9%, 21.8%) | 10.2<br>(8.3, 12.2) | 16.8%<br>(12.3%, 21.9%) |
| Reduction due to TB | 2.4<br>(1.6, 3.3) | 52.8%<br>(41.8%, 62.7%) | 4.5<br>(2.9, 7.0) | 41.6%<br>(29.9%, 52.8%) | 17.0<br>(14.0, 19.3) | 8.0%<br>(5.2%, 11.3%) |
| Reduction compared to prompt treatment | Reference | – | 2.1<br>(1.1, 3.6) | 19.3%<br>(9.7%, 29.2%) | 14.6<br>(12.2, 16.4) | -9.0%<br>(-13.6%, -4.8%) |
| *Alternative specification 2 (no TB disease progression)* |  |  |  |  |  |  |
| Absolute value | 22.4<br>(20.3, 23.8) | 1.5%<br>(-1.9%, 5.0%) | 21.6<br>(19.3, 23.0) | 10.6%<br>(4.1%, 19.3%) | 16.5<br>(14.7, 18.3) | 89.6%<br>(71.8%, 107.8%) |
| Reduction due to TB | 4.8<br>(3.8, 5.7) | 5.6%<br>(-10.4%, 17.9%) | 5.6<br>(4.4, 6.9) | 25.9%<br>(14.1%, 35.7%) | 10.7<br>(8.3, 12.7) | 42.3%<br>(37.0%, 47.5%) |
| Reduction compared to prompt treatment | Reference | – | 0.9<br>(0.5, 1.5) | 66.6%<br>(60.4%, 72.5%) | 5.9<br>(4.3, 7.4) | 56.0%<br>(49.5%, 62.4%) |
| *Alternative specification 3 (no post-TB sequelae and no TB disease progression)* |  |  |  |  |  |  |
| Absolute value | 24.9<br>(22.5, 26.3) | 12.7%<br>(7.6%, 18.0%) | 23.9<br>(21.5, 25.4) | 22.7%<br>(14.1%, 33.1%) | 18.2<br>(16.1, 20.1) | 108.9%<br>(88.8%, 129.6%) |
| Reduction due to TB | 2.3<br>(1.6, 3.0) | 54.7%<br>(41.5%, 65.1%) | 3.3<br>(2.3, 4.5) | 57.1%<br>(47.5%, 65.3%) | 9.0<br>(6.6, 11.1) | 51.4%<br>(44.6%, 58.1%) |
| Reduction compared to prompt treatment | Reference | – | 1.0<br>(0.5, 1.7) | 61.9%<br>(55.6%, 68.1%) | 6.7<br>(5.0, 8.4) | 50.1%<br>(42.7%, 57.0%) |
| **DALYs due to TB** |  |  |  |  |  |  |
| *Main analysis* |  |  |  |  |  |  |
| Absolute value* | 11.4<br>(8.9, 14.2) | 0% | 17.1<br>(13.1, 22.1) | 0% | 37.7<br>(34.3, 40.3) | 0% |
| Reduction compared to prompt treatment | Reference | 0% | 5.7<br>(3.5, 8.6) | 0% | 26.3<br>(23.6, 28.5) | 0% |
| *Alternative specification 1 (no post-TB sequelae)* |  |  |  |  |  |  |
| Absolute value | 5.3<br>(4.1, 7.3) | 51.5%<br>(41.1%, 61.6%) | 9.9<br>(7.0, 14.0) | 42.1%<br>(30.9%, 53.4%) | 34.4<br>(30.8, 37.2) | 9.0%<br>(5.9%, 13.2%) |
| Reduction compared to prompt treatment | Reference | – | 4.4<br>(2.6, 7.0) | 23.0%<br>(12.1%, 36.9%) | 28.8<br>(25.9, 31.4) | -9.5%<br>(-14.9%, -4.6%) |
| *Alternative specification 2 (no TB disease progression)* |  |  |  |  |  |  |
| Absolute value | 10.8<br>(8.6, 12.8) | 5.3%<br>(-11.6%, 18.4%) | 12.5<br>(10.0, 15.0) | 26.3%<br>(14.6%, 35.9%) | 23.5<br>(19.8, 26.6) | 37.9%<br>(32.9%, 43.3%) |
| Reduction compared to prompt treatment | Reference | – | 1.8<br>(1.0, 2.9) | 69.1%<br>(62.1%, 76.5%) | 12.7<br>(10.3, 15.1) | 51.9%<br>(45.4%, 58.1%) |
| *Alternative specification 3 (no post-TB sequelae and no TB disease progression)* |  |  |  |  |  |  |
| Absolute value | 5.3<br>(4.1, 6.6) | 53.7%<br>(40.9%, 64.3%) | 7.3<br>(5.4, 9.6) | 57.2%<br>(47.4%, 66.1%) | 19.8<br>(16.2, 23.3) | 47.5%<br>(41.1%, 53.7%) |
| Reduction compared to prompt treatment | Reference | – | 2.0<br>(1.2, 3.4) | 64.3%<br>(57.5%, 71.7%) | 14.6<br>(11.8, 17.3) | 44.7%<br>(37.1%, 51.8%) |

Estimates of TB-attributable DALYs showed similar changes under the alternative model specifications. Compared to the main analysis, alternative specification 3 (omitting both post-TB sequelae and TB disease progression) produced estimates of TB-attributable DALYs that were 48-57% lower across scenarios, and with the estimated DALYs resulting from delayed or no TB treatment (vs. prompt treatment) reduced by 45-64%.

Figure S3 and S4 report partial rank correlation coefficients (PRCCs) describing the relative sensitivity of results to changes in individual parameters. For total TB DALYs under the prompt treatment scenario, the most influential parameters were the extent of FEV1 recovery following TB cure (PRCC = -0.66), the probability of loss-to-follow-up from ART (PRCC = 0.47), and the HIV-specific mortality rate on ART (PRCC = 0.46). For the increase in DALYs with delayed treatment (vs. prompt treatment), the most influential parameters were the rate of care-seeking for individuals with untreated TB, off ART (PRCC = -0.92), the rate of care-seeking for individuals with untreated TB, on ART (PRCC = -0.86), and the extent of FEV1 recovery following TB cure (PRCC = -0.51).

## Discussion

In this study, we estimated the lifetime health losses caused by TB among individuals attending routine TB diagnosis in settings with high levels of co-prevalent HIV. We estimated these outcomes using a novel mathematical model of TB progression and treatment that incorporates a growing understanding of the progressive lung damage caused by TB, and the health consequences of post-TB sequelae among TB survivors.

Our results demonstrate substantial reductions in life expectancy associated with TB. Even with prompt treatment, individuals with TB had an average reduction in life expectancy of >5 years. Delayed treatment—as may result from a missed TB diagnosis or loss-to-follow-up—increased the loss in life expectancy by approximately 50%, highlighting the urgency of prompt diagnosis treatment initiation, and continuity of care. Estimated DALYs (capturing the impact of TB on both mortality and quality-of-life) showed similar results, and also revealed the extent of health losses associated with post-TB sequelae.

These post-TB health losses were particularly large for individuals with delayed diagnosis, reflecting the additional lung damage associated with a prolonged duration of disease. The magnitude of these post-TB health losses are consistent with recent global estimates,^29^ and empirical estimates for Malawi,^30^ that point to substantial ongoing morbidity and elevated mortality among TB survivors.

The qualitative results of our main analysis were similar to the alternative modelling approaches we examined, with life expectancy shorter (and TB-attributable DALYs greater) with increasing delays to diagnosis, and for individuals with HIV compared to those without HIV. However, the magnitude of these estimates was starkly different, with the results of the alternative specifications (reflecting earlier modelling approaches) suggesting smaller effects of TB on life expectancy and DALYs. These differences in the estimated magnitude of health effects could be consequential for policy analyses, as they suggest that the health gains achieved by preventing TB could be substantially greater than in earlier analyses. Similarly, the health gains produced by interventions that achieve prompt treatment for individuals with symptomatic disease may be greater than previously thought.

This study has several limitations. First, the modelled analyses used to project lifetime health consequences relied on several assumptions (in particular, the long-term causal effects of TB under different treatment scenarios) that are difficult to verify empirically. While randomized assignment of individuals to TB disease is unethical, quasi-experimental studies are starting to generate evidence confirming the magnitude of post-TB health effects.^10,31,32^ These studies are valuable, yet more are needed (and with larger cohorts) to understand how post-TB health consequences vary across settings and under different treatment conditions. Second, this study only examined a small number of scenarios, reflecting the potential impact of delays in TB treatment, or lack of access to treatment. While these results have clear relevance for program strategy, additional analyses will be needed to assess the health impact of interventions designed to increase the timeliness and coverage of TB treatment, taking account of the specific health benefits that can be achieved by the intervention as well as the costs. Moreover, it is possible that new interventions may allow for greater recovery of lung function during TB treatment, or better rehabilitation for TB survivors, reducing the long-term health consequences of TB compared to those estimated in this analysis.^33^ Third, this analysis did not consider the consequences of secondary TB cases resulting from ongoing *Mtb* transmission by individuals with an extended duration of untreated disease. While there is little evidence on the number of secondary cases that result from delayed diagnosis, the inclusion of these outcomes would only increase the health benefits estimated for with earlier treatment initiation. Finally, our analyses were conducted for three settings with high levels of TB and HIV burden, and with notable gaps in healthcare access. Results will likely be different for other settings, and results should be generalized with caution.

Despite these limitations, our analysis demonstrates the implications of new natural history evidence on TB lung damage and post-TB sequelae for the lifetime health consequence caused by TB, and the timeliness of TB diagnosis. Analyses for TB health burden and the impact of TB interventions should take account of these factors to capture the full consequences of TB over the life course of affected individuals.

## Data Availability

All data produced in the present study are available upon reasonable request to the authors.

