## Supplementary materials for "Implications of progressive lung damage and post-TB sequelae for the health benefits of prompt TB diagnosis in high HIV prevalence settings: a mathematical modeling analysis"

### Supplementary exhibits

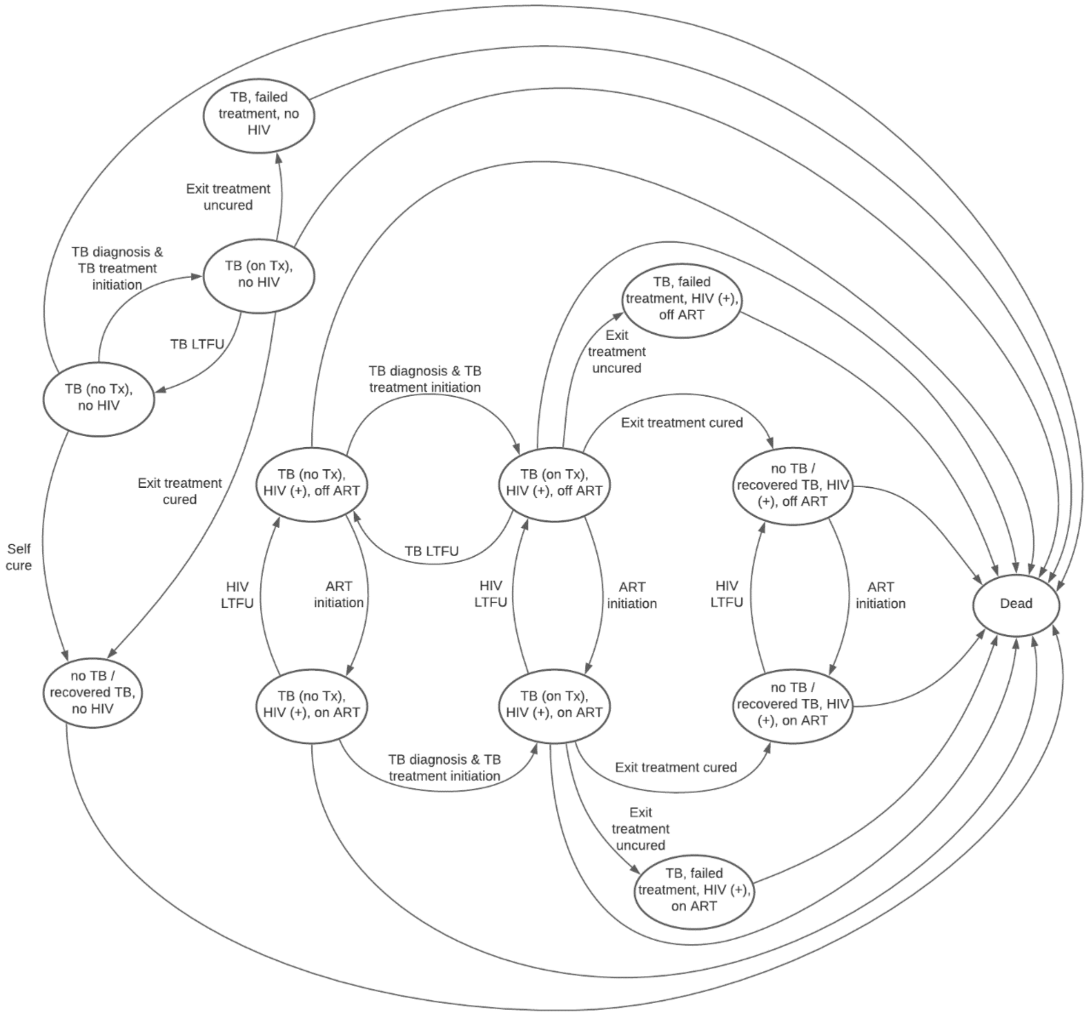

**Figure S1. State transition diagram.**

| Parameter | Value/Assumption (95% interval) | Parameterization for probabilistic sensitivity analysis | Source of value / prior distribution |
|---|---|---|---|
| ***Mortality*** | | | |
| Background (non-TB/non-HIV) annual mortality rate ($\boldsymbol{\mu_{i,j}^{BG}}$) | Country-, age- and sex-specific | Fixed | (1, 2) |
| Rate ratio for TB mortality by year of age (*reference age = 40*) ($\boldsymbol{RR_i^{TB}}$) | Age-specific (see Figure S8) | Fixed | (2) |
| Annual rate of TB-specific mortality ($\boldsymbol{\mu_{base}^{TB}}$), for age = 40 | 0.291 (0.213, 0.398) | Gamma (37.84, 130.05) | (3) |
| Annual mortality rate due to treatment side effects (applied to TB treatment) ($\boldsymbol{\mu^{Tx}}$) | 0.13 (0.80, 2.10) x 1e-3 | Gamma (15.2, 11686) | (4) |
| Rate ratio for mortality due to lung damage ($\boldsymbol{RR_{FEV1}}$) | Fitted log-linear function, with parameter β = 1.753 (1.59, 1.91) (see Figure S9) | Normal (1.753, 0.082) | (5) |
| HIV-specific annual mortality rate, off ART (CD4 cell values are in cells/μL) * ($\boldsymbol{\mu_{CD4,off\ ART}^{HIV}}$) | Fitted log-linear function, with parameters $a_1$ = -0.011 (-0.012, -0.009) $b_1$ = -0.175 (-0.367, 0.017) (see Figure S10A) | Normal (-0.011, 0.0006) Normal (-0.175, 0.098) | (6) |
| HIV-specific annual mortality rate, on ART (CD4 cell values are in cells/μL) * ($\boldsymbol{\mu_{CD4,on\ ART}^{HIV}}$) | Fitted log-linear function, with parameters $a_2$ = -0.0058 (-0.013, 0.0012) $b_2$ = -3.533 (-5.538, -1.53) (see Figure S10B) | Normal (-0.0058, 0.0036) Normal (-3.533, 1.02) | (7) |
| ***Treatment*** | | | |
| Probability of cure with TB treatment completion ($\boldsymbol{p_{cure}}$) | 0.94 (0.91, 0.96) | Beta (322.3, 20.6) | (8) |
| Annual rate of returning to TB care following discontinuation or failure, off ART ($\boldsymbol{r_{return}^{off\ ART}}$) | 2.0 (1.0, 4.0) | Gamma (6.7, 3.3) | Assumption |
| Annual rate of returning to TB care following discontinuation or failure, on ART ($\boldsymbol{r_{return}^{on\ ART}}$) | 4.0 (2.0, 8.0) | Gamma (6.656, 1.664) | Assumption |
| Probability of LTFU prior to TB treatment initiation | 0.14 (0.07, 0.21) | Beta (12.91, 79.30) | Assumption |
| Annual rate of stopping TB care (LTFU) ($\boldsymbol{r_{LTFU}^{TB}}$) | 0.13 (0.10, 0.20) | Gamma (27.1, 208.8) | (9) |
| Probability of cure at LTFU | 0.20 (0.05, 0.35) | Beta (5.1, 20.4) | Assumption |
| TB regimen duration (weeks) | 26 | Fixed | Assumption |
| Annual base rate of self-cure, for untreated active TB ($\boldsymbol{r_{sc_0}}$) | 0.369 (0.290, 0.471) | Gamma (63.688, 172.596) | (3) |
| ***TB Lung Damage*** | | | |
| Mean FEV1 among individuals with symptomatic pulmonary TB | 63.1% (53.1, 73.1) | Beta (55.1, 32.5) | (10-15) |
| Standard deviation of FEV1 among individuals with symptomatic pulmonary TB | 15% | Fixed | (10, 11, 13-15) |
| Rate of FEV1 decline with untreated TB (percentage points per week) | *HIV-negative:* 0.70 (0.55, 0.85) *HIV-positive:* 1.52 (1.20, 2.00) | Gamma (53.8, 35.9) Gamma (83.5, 119.3) | (10-16) |

| Parameter | Value/Assumption (95% interval) | Parameterization for probabilistic sensitivity analysis | Source of value / prior distribution |
|---|---|---|---|
| Rate of FEV1 increase following TB cure, on treatment (*rebound slope*$_1$) (percentage points per week) | 0.44 (0.34, 0.54) | Gamma (61.3, 139.3) | (10, 13-15, 17-20) |
| Rate of FEV1 increase following TB cure, not on treatment (*rebound slope*$_2$) (percentage points per week) | 0.096 (0.066, 0.126) | Gamma (39.2, 407.9) | (15, 18, 21) |
| Maximum possible recovery of FEV1 following TB cure (*p*) (fraction of 100% minus minimum FEV1 value) | 0.42 (0.30, 0.60) | Beta (16.8, 23.3) | (10-15, 18, 21) |
| ***HIV parameters*** | | | |
| Rate of decline in CD4 count without ART (cells/mm$^3$ per week) ($\boldsymbol{d_{CD4}}$) | 1.17 (0.89, 1.56) | Gamma (46.7, 39.9) | (22) |
| Rate of increase in CD4 count with ART (cells/mm$^3$ per week) | Fitted log-linear function, with parameters<br>$\delta_1$ = 247.2 (244.6, 249.8)<br>$\delta_2$ = -6.96 (-7.02, -6.90)<br>(see Figure S12) | Normal (247.2, 1.32)<br>Normal (-6.96, 0.029) | (23) |
| Annual rate of loss to follow-up from ART ($\boldsymbol{r_{LTFU}^{HIV}}$) | 0.064 (0.044, 0.094) | Gamma (25.0, 390.6) | (24) |
| Time since ART initiation for individuals receiving ART at presentation ($\boldsymbol{t_{ART}}$) | Sampled from a uniform distribution between 3 months and 5 years | Uniform (0.25, 5.0) | Assumption |
| ***Disability weights*** | | | |
| Post-TB disability weights, by FEV1 category | *FEV1 ≥ 0.8:*<br>0.005 (0.003, 0.010)<br>*0.8 > FEV1 ≥ 0.5:*<br>0.047 (0.024, 0.071)<br>*0.5 > FEV1:*<br>0.366 (0.275, 0.458) | Beta (7.6, 1517)<br>Beta (14.4, 292.5)<br>Beta (38.4, 66.6) | (25, 26) |
| TB disability weights, by FEV1 category | *FEV1 ≥ 0.8:*<br>0.092 (0.046, 0.138)<br>*0.8 > FEV1 ≥ 0.5:*<br>0.135 (0.101, 0.203)<br>*0.5 > FEV1:*<br>0.453 (0.340, 0.566) | Beta (13.7, 135.1)<br>Beta (23.0, 147.3)<br>Beta (33.1, 40.0) | (25-27) |
| HIV disability weights, by CD4 category (CD4 count values are in cells/μL) | *CD4 ≥ 350, no ART:*<br>0.012 (0.006, 0.023)<br>*350 > CD4 ≥ 200, no ART:*<br>0.274 (0.184, 0.377)<br>*200 > CD4, no ART:*<br>0.582 (0.406, 0.743)<br>*On ART:*<br>0.078 (0.052, 0.111) | Beta (7.4, 607.6)<br>Beta (22.0, 58.4)<br>Beta (18.3, 13.1)<br>Beta (24.5, 289.8) | (27) |
| Disability weight of TB treatment due to a false-positive TB diagnosis | 0.049 (0.031, 0.072) | Beta (20.7, 400.8) | (27) |
| Discount rate | 0.030 | Fixed | Assumption |

**Table S1. Parameter values.**

* HIV-specific mortality rates assumed to exclude mortality due to TB.

| | Uganda | Kenya | South Africa | Overall sample |
|---|---|---|---|---|
| Number of individuals | 10,000 | 10,000 | 10,000 | 30,000 |
| Male (%) | 74.9 | 82.8 | 61.5 | 73.1 |
| Age (mean, IQR) | 39 (29 – 45) | 36 (29 – 42) | 41 (31 – 49) | 38 (30 – 45) |
| Living with HIV (%) | 24.3 | 18.8 | 69.3 | 37.5 |
| Receiving ART, among persons with HIV (%) | 58.8 | 78.5 | 69.5 | 68.7 |
| CD4 cell count (cells/μL; mean, IQR) | 333 (68 – 457) | 178 (53 – 250) | 316 (140 – 471) | 297 (100 – 457) |
| FEV1 (%; mean, IQR) | 63 (52 – 75) | 63 (52 – 75) | 63 (52 – 75) | 63 (52 – 75) |

**Table S2. Description of the cohort after reweighting to match national notifications data, and resampling to produce an analytic sample of 10,000 individuals per country.**

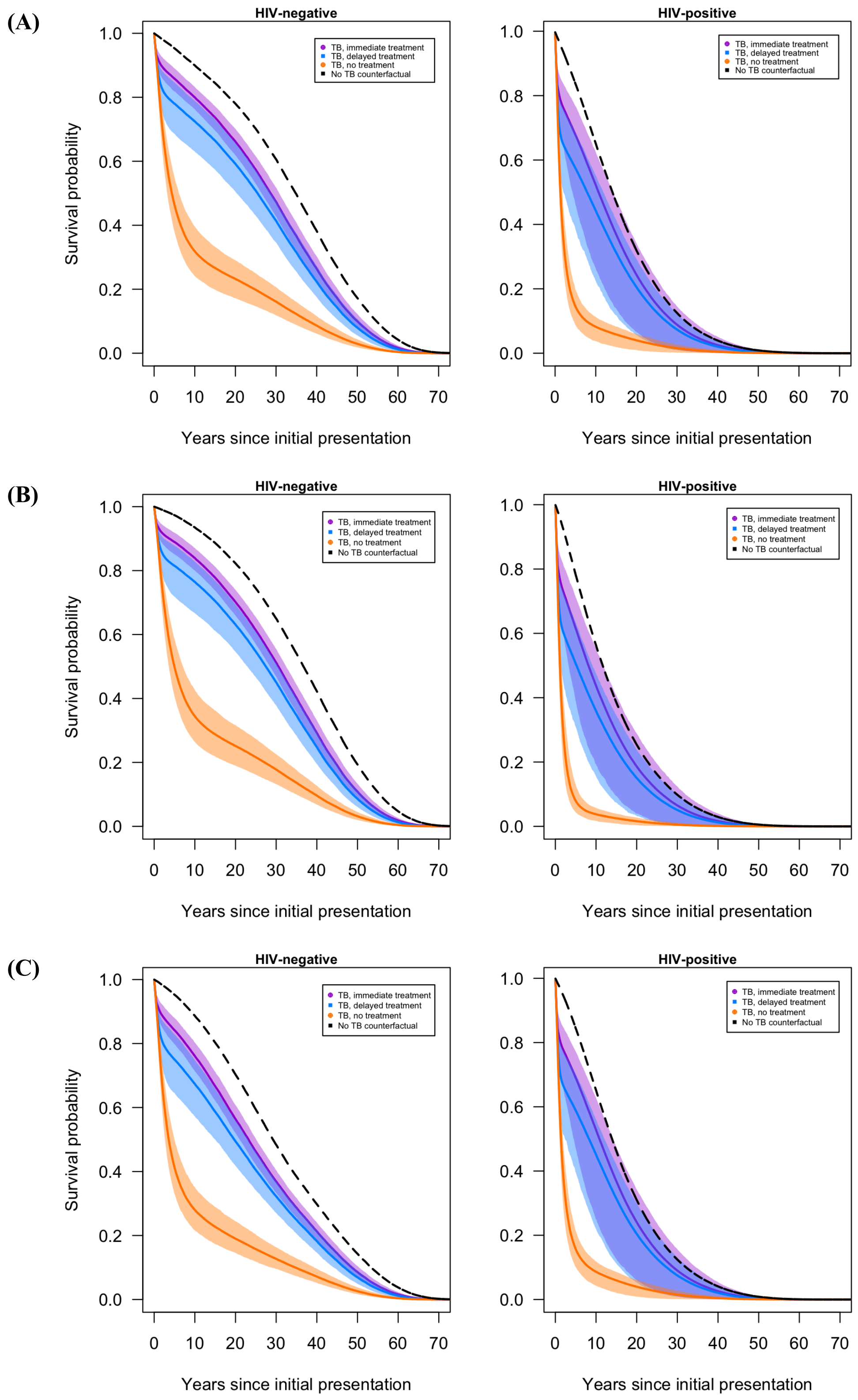

**Figure S2. Survival curves for TB-positive patients in Uganda (Panel A), Kenya (Panel B), and South Africa (Panel C).**

| | HIV-negative | | | HIV-positive | | |
|---|---|---|---|---|---|---|
| | Prompt treatment | Delayed treatment | No treatment | Prompt treatment | Delayed treatment | No treatment |
| ***DALYs due to TB*** | | | | | | |
| Absolute value | 11.7 (8.6, 14.7) | 17.0 (12.2, 23.4) | 35.0 (31.5, 38.3) | 11.1 (7.9, 14.2) | 17.3 (12.1, 23.4) | 39.7 (33.9, 41.9) |
| Difference vs. prompt treatment | Reference | 5.4 (2.7, 9.2) | 23.4 (20.9, 26.1) | Reference | 6.2 (3.2, 10.6) | 28.6 (24.5, 30.9) |
| ***DALYs during TB episode*** | | | | | | |
| Absolute value | 3.7 (2.5, 5.3) | 7.4 (4.4, 12.2) | 30.0 (26.1, 34.0) | 8.3 (5.8, 11.2) | 13.9 (9.3, 20.1) | 38.9 (33.6, 41.2) |
| Difference vs. prompt treatment | Reference | 3.7 (1.7, 7.1) | 26.3 (23.1, 29.6) | Reference | 5.6 (2.8, 9.9) | 30.6 (25.7, 32.7) |
| ***DALYs after TB episode*** | | | | | | |
| Absolute value | 8.0 (5.6, 10.5) | 9.6 (6.8, 13.3) | 5.0 (3.4, 7.3) | 2.8 (1.1, 4.3) | 3.4 (1.4, 5.4) | 0.8 (0.4, 1.4) |
| Difference vs. prompt treatment | Reference | 1.7 (0.9, 3.1) | -3.0 (-4.8, -1.2) | Reference | 0.6 (0.2, 1.3) | -1.9 (-3.1, -0.8) |
| ***YLDs due to TB*** | | | | | | |
| Absolute value | 1.1 (0.6, 1.9) | 1.8 (0.9, 3.6) | 1.2 (0.7, 2.3) | 0.6 (0.3, 1.0) | 1.0 (0.5, 1.8) | 0.4 (0.2, 0.6) |
| Difference vs. prompt treatment | Reference | 0.7 (-1.2, 5.6) | 0.1 (-3.0, 7.1) | Reference | 0.4 (-0.9, 2.9) | -0.2 (-2.1, 2.3) |
| ***YLDs during TB episode*** | | | | | | |
| Absolute value | 0.2 (0.1, 0.2) | 0.3 (0.2, 0.5) | 0.2 (0.1, 0.2) | 0.1 (0.1, 0.2) | 0.2 (0.1, 0.3) | 0.1 (0.1, 0.2) |
| Difference vs. prompt treatment | Reference | 0.2 (-0.1, 1.0) | 0.0 (0.0, 0.0) | Reference | 0.1 (-0.1, 0.6) | 0.0 (0.0, 0.0) |
| ***YLDs after TB episode*** | | | | | | |
| Absolute value | 1.0 (0.5, 1.7) | 1.5 (0.7, 3.2) | 1.0 (0.5, 2.2) | 0.5 (0.2, 0.9) | 0.7 (0.3, 1.6) | 0.2 (0.1, 0.5) |
| Difference vs. prompt treatment | Reference | 0.5 (-1.4, 4.9) | 0.1 (-3.0, 7.1) | Reference | 0.3 (-1.0, 2.4) | -0.2 (-2.1, 2.3) |
| ***YLLs due to TB*** | | | | | | |
| Absolute value | 10.5 (7.9, 13.1) | 15.2 (11.1, 20.2) | 33.9 (30.4, 37.0) | 10.5 (7.5, 13.5) | 16.3 (11.4, 22.3) | 39.3 (33.7, 41.5) |
| Difference vs. prompt treatment | Reference | 4.7 (-0.1, 42.9) | 23.3 (-0.1, 53.9) | Reference | 5.8 (-0.2, 50.6) | 28.8 (0.0, 55.8) |
| ***YLLs during TB episode*** | | | | | | |
| Absolute value | 3.5 (2.5, 5.3) | 7.1 (4.1, 11.7) | 29.9 (25.9, 33.9) | 8.2 (5.7, 11.1) | 13.7 (9.2, 19.8) | 38.7 (33.5, 41.2) |
| Difference vs. prompt treatment | Reference | 3.5 (0.0, 50.4) | 26.3 (0.0, 59.6) | Reference | 5.5 (0.0, 52.5) | 30.6 (0.0, 56.9) |
| ***YLLs after TB episode*** | | | | | | |
| Absolute value | 7.0 (5.0, 9.0) | 8.2 (6.0, 10.4) | 4.0 (2.8, 5.3) | 2.3 (0.9, 3.5) | 2.6 (1.1, 4.0) | 0.6 (0.3, 0.9) |
| Difference vs. prompt treatment | Reference | 1.2 (-9.6, 11.3) | -3.0 (-19.0, 14.0) | Reference | 0.4 (-5.7, 6.9) | -1.7 (-13.1, 3.4) |

**Table S3. Disability-adjusted life years (DALYs), years lost due to disability (YLDs), and years of life lost (YLLs) under prompt, delayed, and no treatment scenarios, stratified by HIV status.**

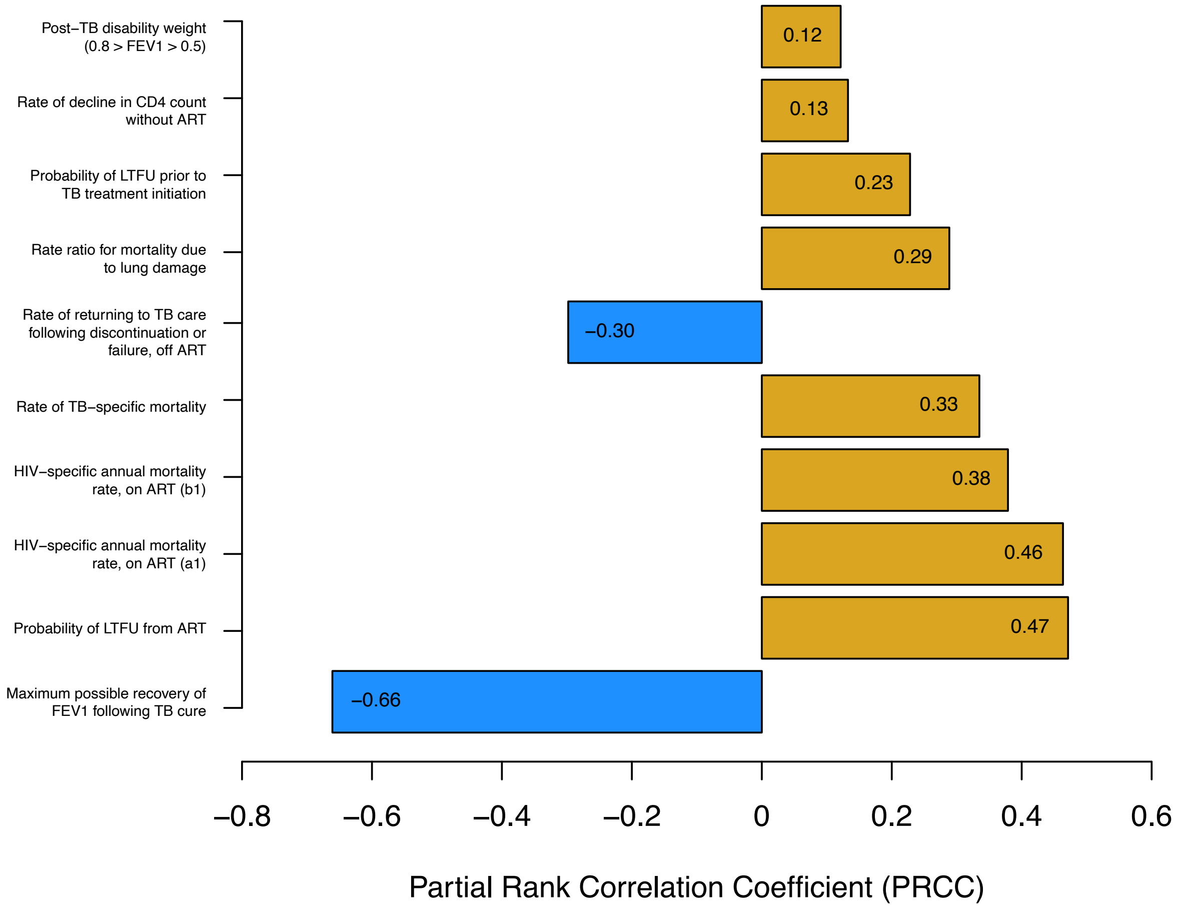

**Figure S3. Partial rank correlation coefficients (PRCCs) for ten parameters with greatest influence on the estimated DALYs due to TB disease with prompt treatment.**

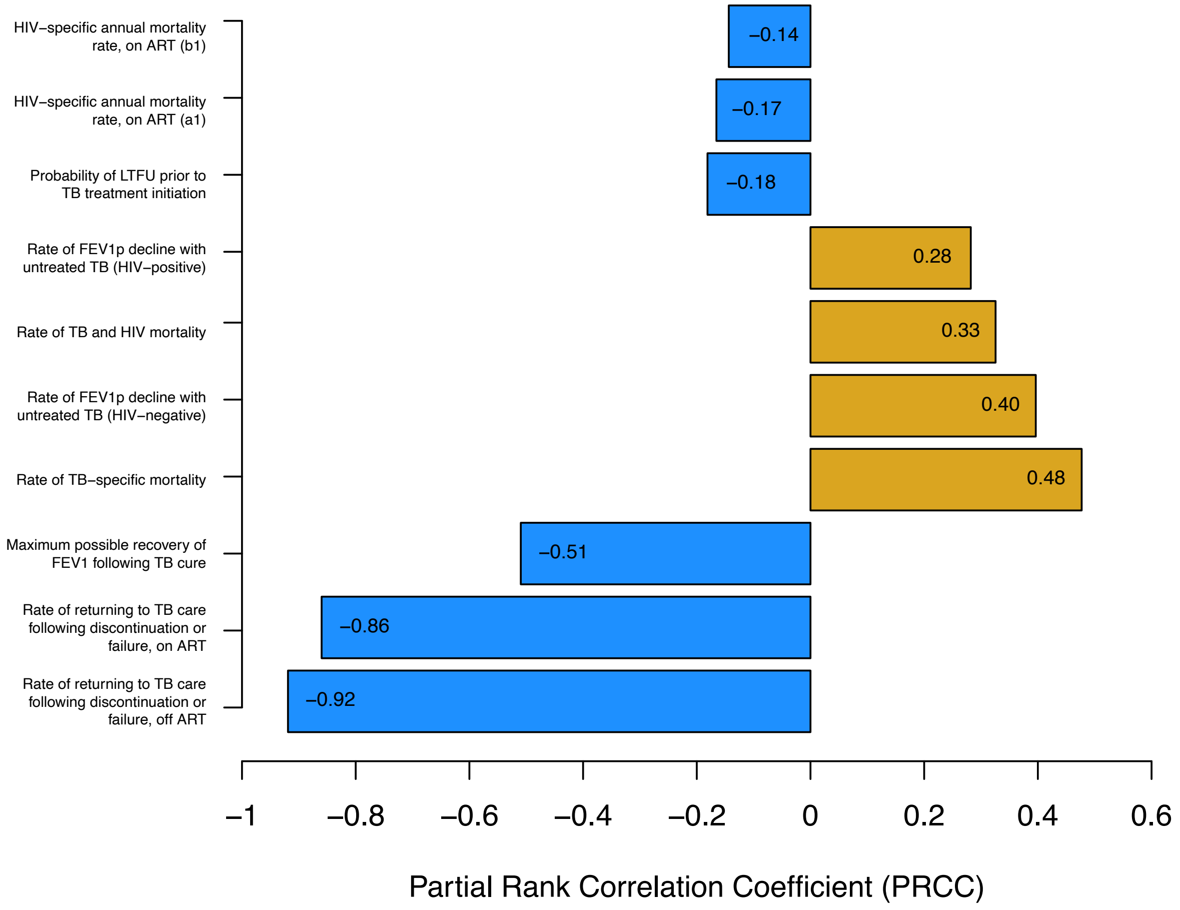

**Figure S4. Partial rank correlation coefficients (PRCCs) for ten parameters with greatest influence on the increase in DALYs due to TB disease resulting from delayed treatment (vs. prompt treatment).**

### Supplementary methods: study cohort

The study cohort represented individuals with symptomatic pulmonary TB presenting for diagnosis at routine healthcare settings in one of three countries with a high burden of TB and HIV: Uganda, Kenya, and South Africa. Data to create the study population were drawn from a diagnostic accuracy study conducted in Uganda, Kenya, and South Africa (28). This sample included ≥ 18-year-old primary healthcare attendees with a clinical suspicion of pulmonary TB (defined as individuals who report cough of ≥ 2 weeks plus at least one other typical symptom of TB), excluding individuals who received any anti-TB treatment within the prior 6 months. For participants determined to have TB via sputum culture, we extracted data on age, sex, HIV status, and CD4 cell count if HIV positive. This sample included 400 participants for each of South Africa and Uganda, and 399 for Kenya. Figure S5 shows the sex and age distribution of sampled participants.

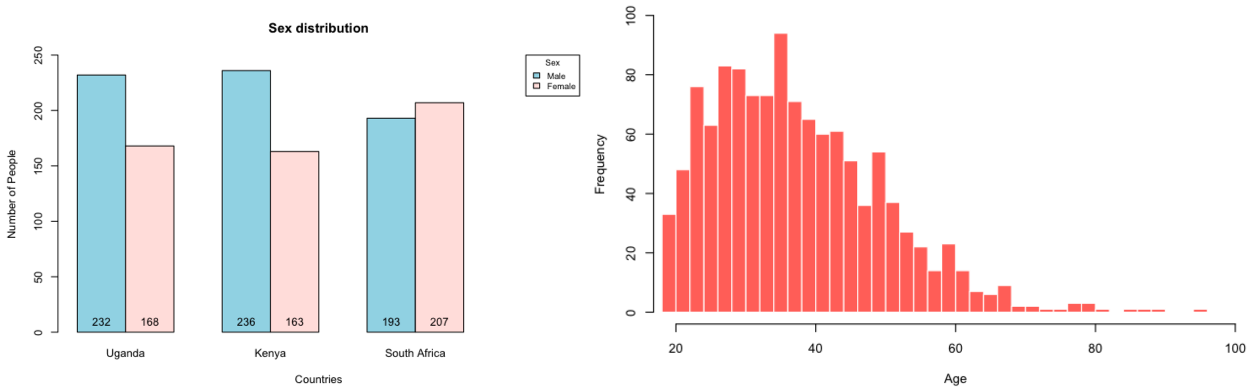

(A) (B)

**Figure S5. Distribution of the study sample by sex (Panel A) and age (Panel B).**

Participants were tested for HIV (unless HIV status was already known), and information was collected on whether participants were currently receiving ART. Figure S6 shows the HIV status of the starting population, and the ART treatment status for those that were HIV-positive. For HIV-positive participants, a CD4 count testing was performed at baseline, if they did not have a documented for CD4 count within the preceding 90 days. Figure S7 shows the distribution of CD4 count values at baseline (average value 400 cells/µL).

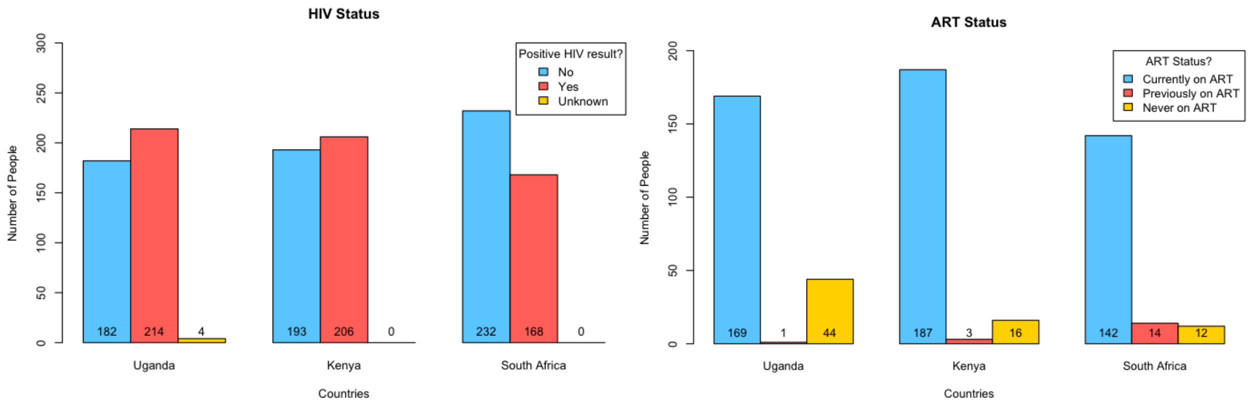

(A) (B)

**Figure S6. Distribution of the study sample by HIV status (Panel A), and ART status among HIV-positive participants (Panel B).**

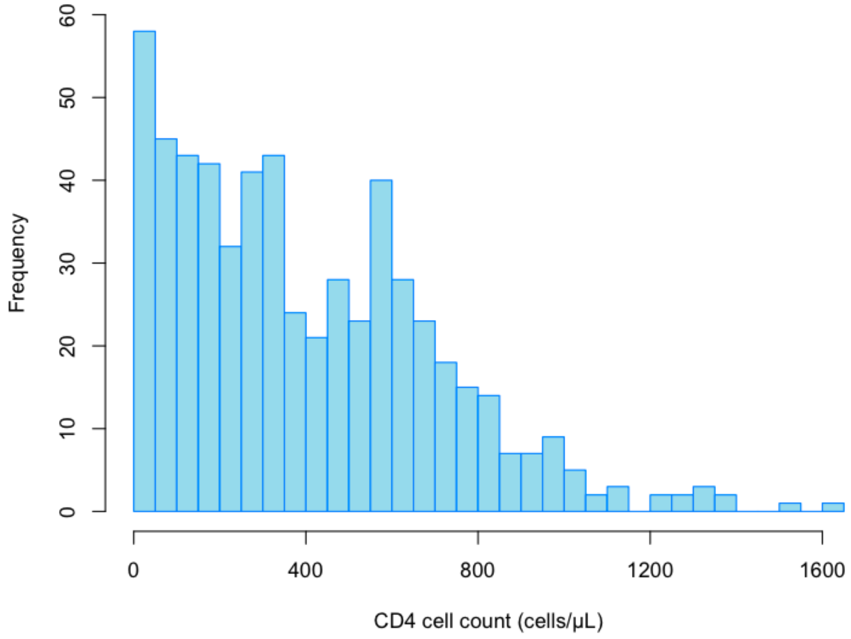

**Figure S7. Distribution of baseline CD4 cell count values among HIV-positive individuals.**

#### Reweighting to match national notifications data

We used iterative proportional fitting (raking) to reweight the study population to match national notifications data in terms of the marginal distributions for age, sex, and HIV status (29). To do so, age was categorized into 6 age groups: 20-24, 25-34, 35-44, 45-54, 55-64, and 65+ years old. Sex was categorized as “male” or “female”. HIV status was categorized as “HIV negative” or “HIV positive”.

After reweighting we resampled the participant data with replacement to create a final analytic cohort of 10,000 individuals per country. Table S2 provides descriptive information on the final analytic sample.

**FEV1 at presentation**

As the data used to create the analytic cohort did not include FEV1, we assigned FEV1 values to participants using evidence from published studies describing the distribution of FEV1 among individuals with diagnosed TB, as described below.

Ralph *et al*. (10) conducted a study among 200 smear-positive pulmonary TB patients in Indonesia. The findings were compared with a control group that consisted of 40 healthy volunteers. At baseline, the mean FEV1 was measured as 63% with a standard deviation of 19.4. In the control group, this mean FEV1 was found to be 92% with a standard deviation of 19.9.

Guessogo *et al*. (11) conducted a study among 28 smear-positive pulmonary TB patients in Cameroon. The authors measured the pulmonary functions of the newly diagnosed TB patients at the time of diagnosis (baseline) and two months after treatment initiation. They compared the results with 19 healthy subjects. In this study, the mean value for baseline FEV1 among TB patients was measured as 1.8L with a standard deviation of 0.7. The FEV1 value among healthy subjects was measured as 3.0L with a standard deviation of 0.8. With the assumption of considering FEV1 value among healthy subjects as 100%, the baseline FEV1 among TB patients was calculated as 60% with a standard deviation of 23.3.

Maguire *et al*. (12) assessed the pulmonary function of smear-positive pulmonary TB patients in Indonesia over 6 months. Spirometry values at the time of the diagnosis had a mean of 66.9% with a standard deviation of 2.7, among patients who completed all visits.

Radovic *et al*. (13) conducted a study among 40 patients newly diagnosed with extensive pulmonary TB in Serbia. At the beginning of the treatment, the mean of FEV1 value was measured as 73.5% with a standard deviation of 16.3.

Chesov *et al*. (14) investigated the pulmonary function of patients at the time of TB diagnosis before TB treatment initiation and at the end of TB treatment. This study was conducted among 278 pulmonary TB patients in Ukraine. Prior to starting TB treatment, the mean FEV1 value was found to be 56.0% with a standard deviation of 22.0.

Allwood *et al*. (15) conducted a study to observe the changes in pulmonary function of newly diagnosed TB patients over the course of their TB episode. They measured the spirometry values of 43 patients at the time of diagnosis, and several follow-up visits: 2 and 6 months after treatment initiation, 6 and 12 months after treatment completion. The median of FEV1 is found to be 59% (IQR: 50%–77%) at the time of diagnosis.

The central estimate from these different studies was a FEV1 at the point of TB diagnosis of 63.1, calculated as the simple mean across studies. We created an uncertainty interval for this mean by assuming the duration of the disease progression until treatment initiation is uniformly distributed. We used the standard deviations reported by each empirical study to parameterize the individual level distribution of values around this overall mean, assuming they followed a Beta distribution.

### Supplementary methods: modelling approach for TB natural history

#### Mortality

Mortality rates were calculated using an excess mortality model, with an individual's total mortality rate calculated as the sum of an age-specific and sex-specific background mortality rate and disease-specific mortality rates. Disease-specific mortality rates include components representing the additional mortality associated with lung damage, prevalent TB disease, and HIV (among HIV-positive individuals).

Background mortality rates

Background mortality rates ($\mu^{BG}$) were based on cause-deleted life tables, to avoid double counting of HIV and TB mortality. Cause-deleted life tables (by country and sex) were created by multiplying all-cause mortality rates for each year of age by 1 minus the proportion of total mortality for that age that was attributed to TB or HIV. All-cause mortality rates for each country and sex were extracted from the United Nations World Population Prospects 2019 revision (1). Estimates of the proportion of total mortality attributable to TB and/or HIV were extracted from the Global Burden of Disease (GBD) 2019 study (2).

TB-specific mortality rate

The TB-specific mortality rate ($\mu^{TB}$) (i.e., additional mortality during the TB episode on top of the contribution from lung damage described above) was calculated as a function of age and FEV1. Figure S8 shows how the TB-specific mortality rate varied by age. These estimates were calculated using estimates of TB cases and deaths reported by the Global Burden of Disease (GBD) 2019 study (2) by age group, and using spline interpolating to obtain values by single year of age.

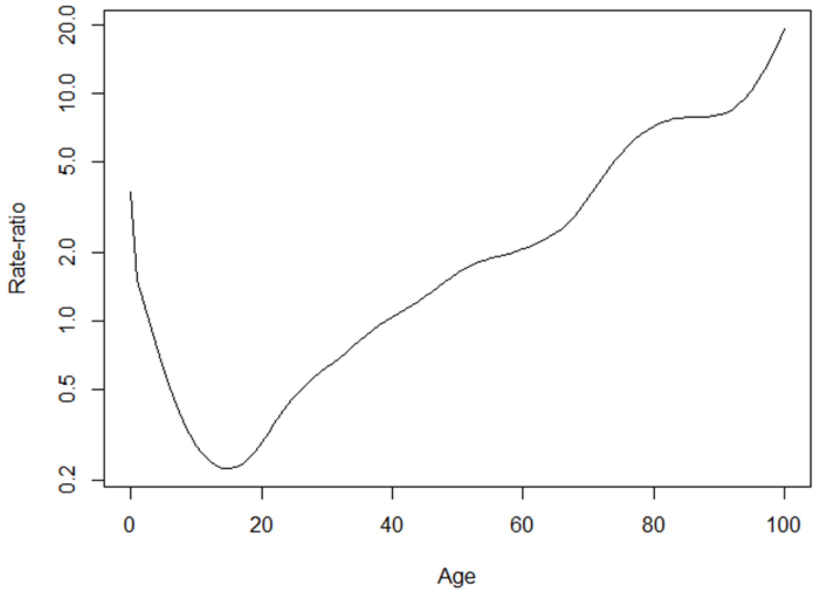

**Figure S8. TB mortality rate by year of age (relative to a 40-year-old individual).**

Mortality rate due to lung damage

We modelled additional mortality for individuals with TB-associated lung damage, as measured by FEV1. We parameterize this component using data from Duong *et al*. (5), which reports mortality rate ratios for individuals with different levels of lung function as represented by different FEV1 ranges: no impairment, mild impairment, moderate impairment, severe impairment. We assumed that the mortality rate ratio increases log-linearly as a function of FEV1 for individuals with an FEV1 below 100, as shown in Figure S9. By fitting the log-linear function to the data in (5), we obtained an estimate for the parameter $\beta$ of 1.753, with a standard deviation of 0.082.

$$RR_{FEV1} = \exp(\alpha + \beta \times (1 - FEV1p)) \tag{1}$$

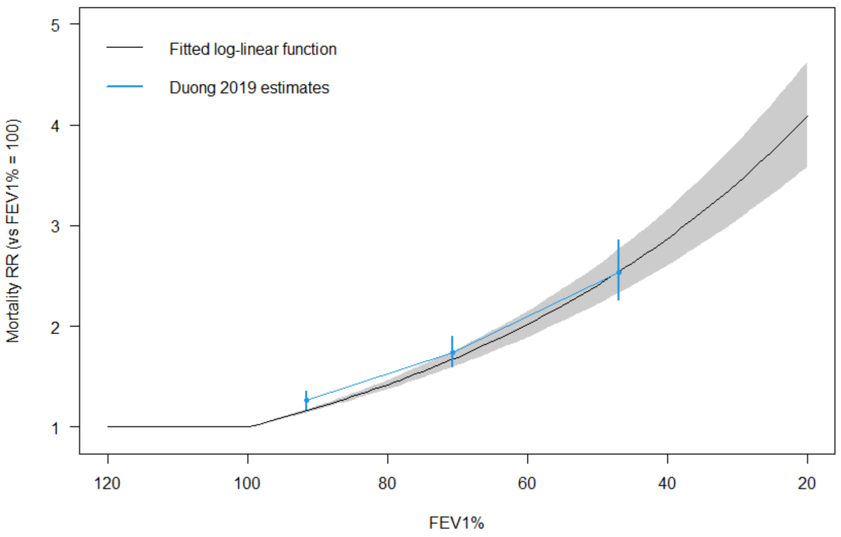

**Figure S9. Mortality rate ratios for FEV1.**

The additional mortality rate for individuals with TB-associated lung damage was applied to individuals with prevalent TB as well as to TB survivors, as a function of current FEV1.

Mortality rates for individuals with co-prevalent TB and HIV were assumed to be higher than the sum of TB- and HIV-specific rates, based on the very high mortality rates observed for individuals with both diseases.

Equation 2 shows how the overall TB-specific mortality rate was constructed, as a function of a base rate, rate ratio for differences by year of age (Figure S9), the additional TB mortality for individuals with HIV (scaled by normalized CD4 cell count value), and finally a scaling of this overall rate by the extent of lung damage, represented by FEV1.

$$\mu_{i,FEV1,CD4}^{TB} = \left(\mu_{base}^{TB} * RR_i^{TB} + \mu^{TB-HIV} * \left(1 - min\left(CD4,\ CD4_{ref}\right)/CD4_{ref}\right)\right) * (1 - FEV1p) \qquad (2)$$

HIV-specific mortality rate

The HIV-specific mortality rate was calculated as a log-linear function of CD4 cell count and ART status (Equation 3). Since the effect of CD4 cell count on the mortality rates would be different for the cases being on ART and off ART, the functions for these two cases are considered separately.

$$\mu_{CD4,ART}^{HIV} = \begin{cases} \exp(a_1 + b_1 \times CD4), if\ ART = F \\ \exp(a_2 + b_2 \times CD4), if\ ART = T \end{cases} \qquad (3)$$

For individuals not receiving ART, the HIV-specific mortality rate was fitted to the reported estimates of untreated HIV mortality in Côte d'Ivoire (6) to obtain the parameters $a_1, b_1$, as shown in Equation 3. Similarly, for individuals receiving ART, the HIV-specific mortality rate was fitted to observational data reported in a multi-country prospective cohort study in sub-Saharan Africa (7), to obtain the parameters $a_2, b_2$. As reported HIV mortality will include TB deaths among individuals with HIV, we assumed that 25% of the reported HIV mortality rate was due to TB deaths, and subtracted that proportion from the HIV-specific mortality rate. The mortality rates corresponding to different CD4 cell count values for those who are not on ART and received ART, respectively, are shown in Figure S10.

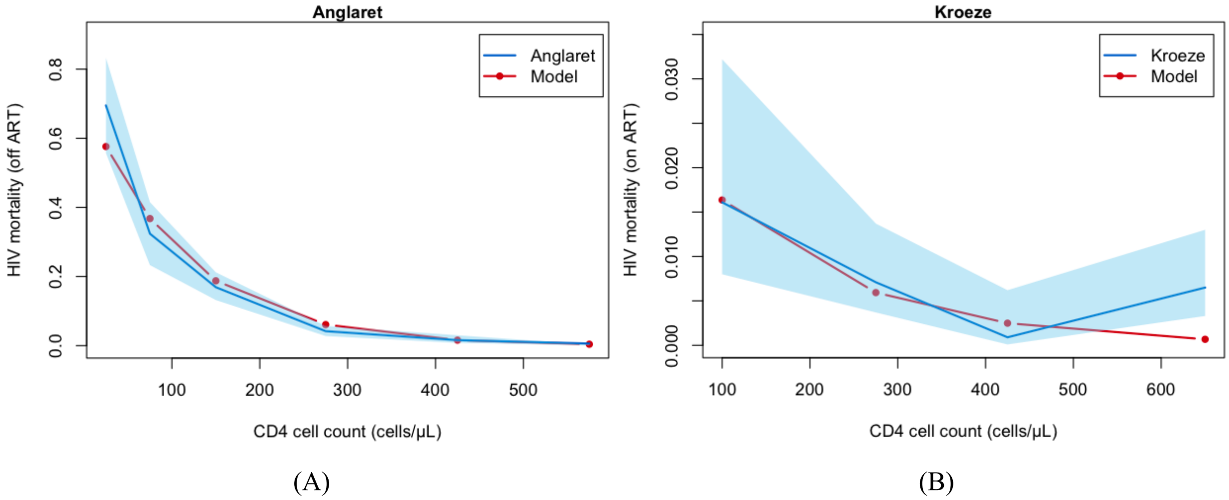

**Figure S10. HIV-specific mortality rate for individuals not receiving ART (Panel A), and receiving ART (Panel B).**

For each week in the model, the probability of death was calculated from the summed mortality rates (Equation 4), where *i* represents the age group and *j* represents sex.

$$p_{mort,t} = 1 - exp\left[-\left(\mu_{i,j}^{BG} * RR_{FEV1} + \mu_{i,FEV1,CD4}^{TB} + \mu_{CD4,ART}^{HIV}\right) * \frac{1}{52}\right] \quad (4)$$

**TB lung damage**

To measure lung damage due to TB disease, we used the FEV1 percentage (‘FEV1’), defined as a person’s expired lung volume in the first second of forced expiration, as compared to average values in the population. In the model, FEV1 was operationalized as an individual-level characteristic updated on a weekly basis.

Parameter values and data sources

In the model, the FEV1 parameter influences a number of other model mechanisms and health outcomes, including TB-specific mortality rates, care-seeking behavior, the rate of self-cure, and disability weights. These relationships are described in other sections.

Figure S11 summarizes assumptions made about the changes in FEV1 over the course of the TB episode (from incident TB to post-TB).

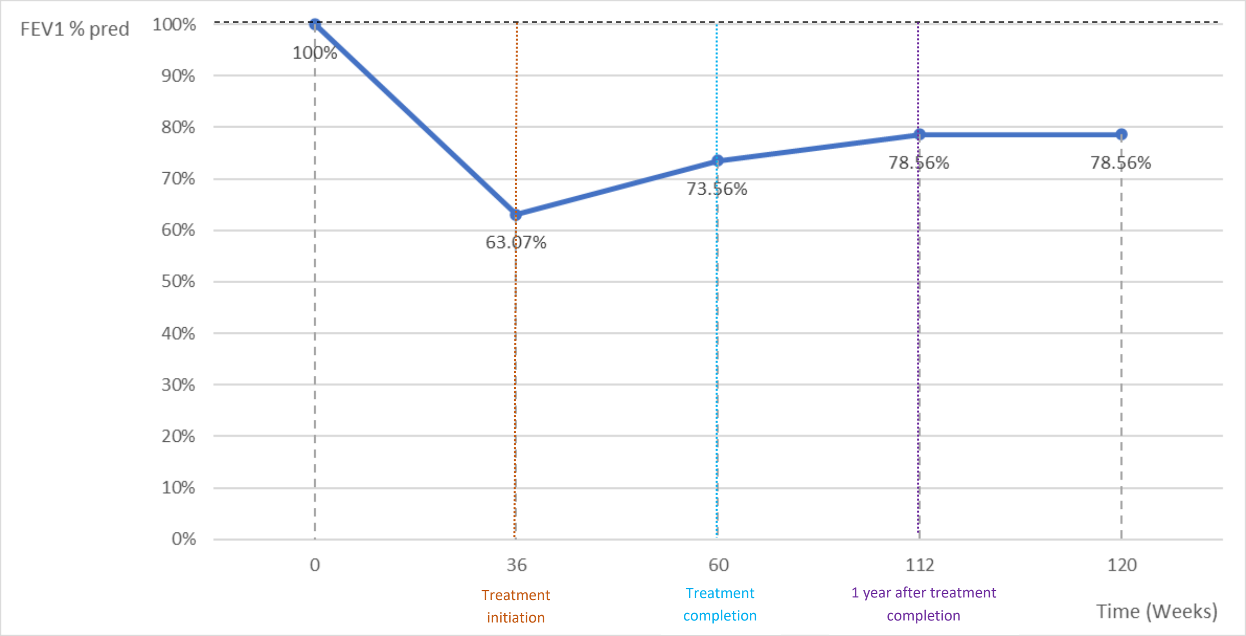

**Figure S11. The changes in FEV1 over the course of the TB episode.**

We made several assumptions to describe the dynamics of FEV1 before, during, and after TB treatment: initial (pre-TB) FEV1 was assumed to be 100%, the duration of the disease progression until treatment initiation was assumed to be uniformly distributed, based on assumptions described in the Global Tuberculosis Report (16). For notified HIV-negative TB cases, the duration of disease was uniformly distributed between 0.2 and 2 years (average 13.2 months). For notified HIV-positive TB cases, the duration of disease was uniformly distributed between 0.01 and 1 years (average 6.0 months). We assumed the duration of TB treatment would be 6 months.

*The weekly rate of FEV1 decline for individuals with untreated TB disease*

This rate was calculated by using estimates of FEV1 at the time of TB diagnosis and duration of untreated disease, assuming that FEV1 was 100% for individuals newly developing TB. The duration of disease was assumed to be uniformly distributed over the interval (0.2, 2) years and (0.01, 1) years for HIV-negative and HIV-positive individuals with TB, respectively (16). The rate of decline was estimated by dividing the difference between the initial FEV1 value and the FEV1 at presentation by the duration of disease (assuming a linear rate of decline). The median estimate for this rate of decline was 0.70 percentage points per week for HIV-negative individuals, and 1.52 percentage points per week for HIV-positive individuals.

*The rate of improvement for individuals receiving TB treatment (rebound $slope_1$)*

For this parameter, we compared estimates of FEV1 at treatment initiation to FEV1 values measured at treatment completion and 24 weeks/26 weeks or 6 months after initiating the treatment.

In Ralph *et al*. (10), changes in FEV1 were assessed during treatment, with FEV1 value after 6 months estimated as 71% with a standard deviation of 17.5.

In Radovic *et al*. (13), FEV1 was assessed at 2 and 6 months after initiation the treatment. The mean FEV1 value at treatment completion was measured as 72.0%, with a standard deviation of 20.1.

In Chesov *et al*. (14), the mean FEV1 value for TB patients with a positive treatment outcome ($n$ = 206) was 83.2% with a standard deviation of 22.6.

In Allwood *et al*. (15), the treatment completion is assumed to happen 6 months after initiating the treatment. At that time point, the median FEV1 value is found to be 79% with an interquartile range of 0.59–0.87.

Khosa *et al*. (17) assessed the spirometric values at 8, 26, and 52 weeks after initiation of TB treatment among 62 patients in Mozambique. At 26 weeks FEV1 was 71.4% with a standard deviation of 17.7.

Meghji *et al*. (18) investigated the lung damage and associated outcomes among patients who completed TB treatment in Malawi. The mean FEV1 value at the time of treatment completion was calculated as 74.8%.

Pefura-Yone *et al*. (20) evaluated the relationship between TB treatment completion and chronic respiratory symptoms based on spirometry measures among 177 patients in Cameroon. The mean FEV1 value at the end of TB treatment was measured as 76.3% with a standard deviation of 19.0.

Using the average of the values from these studies, the rebound $slope_1$ (weekly improvement in FEV1 during treatment) was estimated to be 0.44 percentage points per week.

*FEV1 improvement after treatment (rebound $slope_2$)*

Evidence suggests that FEV1 continues to rebound after treatment completion. For this parameter, we compared FEV1 values measured at treatment completion versus 1 year after initiating the treatment. Nightingale *et al*. (30) evaluated the changes in lung functions of the patients who completed TB treatment over 3 years. The authors found that most of the TB patients showed the maximum improvement in their FEV1 values in the first year after treatment completion.

In Allwood *et al*. (15), the authors measured changes in FEV1 values in several follow-up visits subsequent to TB treatment initiation, and the study was concluded with the last measurement at 12 months after treatment completion. At that follow-up visit, the FEV1 was measured as 79% with the interquartile range of 64%–89%.

In Meghji *et al*. (18), the second follow-up visit to perform spirometry measurements was done 12 months after completing the treatment. Since the FEV1 parameters were given as z-score, the reference parameter values for FEV1 in (19) was used to calculate the predicted FEV1 value for this time point. As a result, the mean FEV1 value 12 months after TB treatment completion was found to be 78.3%.

Nishi *et al*. (21) analyzed TB patients who completed treatment to assess the progression in their lung function, in Brazil. They collected data at two time points: follow-up 1 (within 1 year after the treatment) ($n$ = 55) and follow-up 2 (one and two years after the first follow-up) ($n$ = 29). The spirometry results at the time of the first follow-up visit were considered as equivalent to 1 year after treatment completion. The mean FEV1 value was 78.4% with a standard deviation 17.5.

Using the average of the values from these three studies, rebound $slope_2$ (weekly improvement in FEV1 in the year after treatment) was found to be 0.096 percentage points per week.

<u>*Maximal recovery of initial FEV1 (p)*</u>

We assumed that, with successful treatment, individuals would be able to recover a fraction of their maximal lung damage (the difference between initial FEV1 and lowest recorded FEV1). This parameter was defined as the extent of FEV1 recovery, as shown in Equation 5:

$$Final\ FEV1\ value = p * \max FEV1 + (1-p) * \min FEV1 \tag{5}$$

This final FEV1 value represented a value that FEV1 would plateau at following TB treatment and cured. The average fractional recovery of FEV1 ($p$) was based on values found in the literature, and was calculated as 0.42 (equivalent to 42% recovery of the pre-TB FEV1, as compared to the total decline in FEV1 due to TB).

#### TB self-cure

We assumed that some individuals with TB disease would self-cure in the absence of treatment. The self-cure rate ($r_{SC}$) was operationalized as a function of CD4 count and FEV1, under the assumption that self-cure rates would be lower with advanced HIV, and lower for individuals with extensive disease. Equation 6 shows the equation used to calculate $r_{SC}$. In this equation, the CD4 reference value ($CD4_{ref}$) was set to 700 cells/µL (assumed to represent complete immunity).

$$r_{SC} = r_{sc_0} * FEV1 * min(CD4,\ CD4_{ref})/CD4_{ref} \quad (6)$$

#### HIV progression

The model subdivides the analytic cohort by HIV status (HIV positive, HIV negative). For simplicity, we assumed there would be no risk of new HIV infection over the simulation period. We assumed that individuals diagnosed with HIV as part of the evaluation for TB would be initiated on ART immediately.

HIV disease progression was tracked by CD4 cell count. For HIV-positive individuals not currently receiving ART (for example, among those lost to follow-up from ART), we assumed that CD4 cell count would decline linearly, to a minimum value of zero. The rate of CD4 cell count decline ($d_{CD4}$) was set to 61 (95% CI: 46–81) cells/mm$^3$ per year, which is equivalent to 1.17 (95% CI: 0.89–1.56) cells/mm$^3$ per week (Equation 7).

$$CD4 = \begin{cases} \max(CD4\ base - d_{CD4}, 0), & \text{if ART} = \text{F} \\ CD4\ base + \delta_1(1 - \exp(-\exp(\delta_2 \times t_{ART})), & \text{if ART} = \text{T} \end{cases} \quad (7)$$

For HIV-positive individuals initiated on ART, we assumed that CD4 count would increase, with the rate of increase declining over time since ART initiation (Figure S12) (22, 31). In Equation 7, $\delta_1$ and $\delta_2$ are the parameters that govern the shape of the function, $t_{ART}$ represents time since ART initiation, and $CD4\ base$ is the baseline CD4 cell count of the individual when they initiated ART.

#### ART loss to follow-up

We allowed patients to be lost to follow-up from ART in the model. Geng *et al.* (24) traced the patients initiating ART in several health clinics in Uganda, Kenya, and Tanzania for >2 years, accounting for individuals who re-initiated ART at other clinics ('self-transfers'). Based on this study, we calculated the mean annual loss to follow-up (LTFU) rate as 0.064 per year.

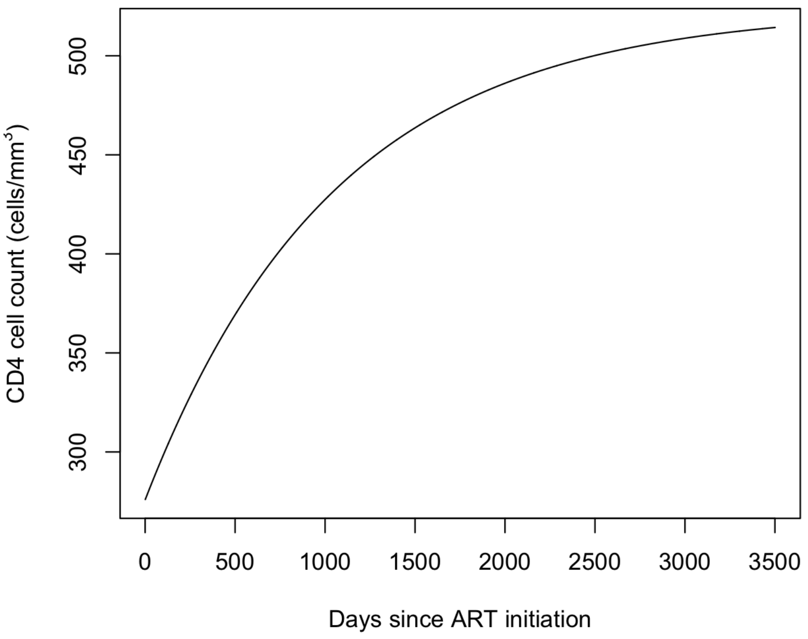

**Figure S12. CD4 trajectory for an individual newly initiated on ART.**

### Supplementary methods: modeling approach for TB treatment

#### TB diagnosis and treatment initiation

The outcomes of TB diagnosis (either true positive or false negative diagnosis) were defined as part of the analytic scenarios, with the 'prompt treatment' scenario representing the outcome of true positive diagnosis and the 'delayed treatment' scenario representing the outcome of false negative diagnosis. For all individuals correctly diagnosed with TB, we assumed there would be a 2-week delay between diagnosis and successful treatment initiation.

#### Treatment completion

Individuals treated for TB were assumed to complete treatment a fixed time since initiation (assuming they had not discontinued before this point), assuming a standard 6-month 1$^{st}$ line treatment regimen.

We assume that the patients could be cured in their first 2 months of treatment. Over the next 4 months, the probability of cure/treatment success conditional on treatment completion ($p$) was assumed to be 0.94 (8).

#### Treatment discontinuation

We assumed that would discontinue treatment (loss to follow-up) at a fixed rate, with these rates calculated to reproduce treatment outcomes data for each country reported in the WHO Global TB database (9). We assumed that a fraction of individuals lost to follow-up would be cured, if they default after the first two months of treatment. This fraction was assumed to be 20%.

For individuals receiving both ART and TB treatment, we assumed that loss to follow-up from one service would imply loss to follow-up from the other. For this purpose, we considered the probability of loss to follow-up as the maximum of the TB treatment and ART loss to follow-up probabilities.

#### Return to care

Among individuals with untreated TB disease, we assumed a fixed rate of return for care. This rate applied to individuals lost to follow-up during diagnosis, receiving a false-negative TB diagnosis, primary default, lost to follow-up during treatment, and those who completed treatment but fail to achieve cure. We assumed the annual rate of return to care for patients who do not receive ART to be 2.0. For individuals receiving ART, we assumed that their rate of return would be 4.0.

### References

1. United Nations. World Population Prospects 2019. In: Department of Economic and Social Affairs, editor. 2019.

2. Institute for Health Metrics Evaluation. Global Burden of Disease Collaborative Network. Global Burden of Disease Study 2019 (GBD 2019) Results. Seattle, United States: Institute for Health Metrics and Evaluation; 2020.

9. World Health Organization. Case Notifications. Geneva; 2020.

16. World Health Organization. Global Tuberculosis Report 2013. Geneva: World Health Organization; 2013.

17. Khosa C, Bhatt N, Massango I, Azam K, Saathoff E, Bakuli A, et al. Development of chronic lung impairment in Mozambican TB patients and associated risks. BMC pulmonary medicine. 2020;20(1):1-11.

18. Meghji J, Lesosky M, Joekes E, Banda P, Rylance J, Gordon S, et al. Patient outcomes associated with post-tuberculosis lung damage in Malawi: a prospective cohort study. Thorax. 2020;75(3):269-78.

19. Pasipanodya JG, Miller TL, Vecino M, Munguia G, Garmon R, Bae S, et al. Pulmonary impairment after tuberculosis. Chest. 2007;131(6):1817-24.

20. Pefura-Yone EW, Kengne AP, Tagne-Kamdem PE, Afane-Ze E. Clinical significance of low forced expiratory flow between 25% and 75% of vital capacity following treated pulmonary tuberculosis: a cross-sectional study. BMJ open. 2014;4(7):e005361.

21. Nishi MP, Mancuzo EV, Sulmonett N, Almeida INd, César ALA, Miranda SSd. Pulmonary functional assessment: longitudinal study after treatment of pulmonary tuberculosis. Revista do Instituto de Medicina Tropical de São Paulo. 2021;63.
